# Implementation outcomes of tuberculosis digital adherence technologies: a scoping review using the RE-AIM framework

**DOI:** 10.1101/2024.06.11.24308660

**Authors:** Chimweta I Chilala, Nicola Foster, Shruti Bahukudumbi, Mona S. Mohamed, Miranda Zary, Cedric Kafie, Barbie Patel, Genevieve Gore, Kevin Schwartzman, Ramnath Subbaraman, Katherine Fielding

## Abstract

**Introduction:** Globally, tuberculosis (TB) remains one of the leading infectious causes of death, with 1.3 million deaths. Digital adherence technologies (DATs) have the potential to provide person-centred care and improve outcomes. Using the RE-AIM framework, we conducted a scoping review of DAT implementations for TB treatment.

**Methods:** We searched eight databases for papers published between January 2000 and April 2023, using keywords for ‘tuberculosis’ and ‘digital adherence technology’. Articles meeting prespecified inclusion criteria and containing data on RE-AIM domains were included (PROSPERO-CRD42022326968). We defined ‘reach’ as comprising cellphone ownership and engagement by people with TB (PWTB) with DATs, ‘adoption’ as engagement by healthcare providers with DAT programs, ‘implementation’ as the fidelity of the DAT program implemented, and ‘maintenance’ as longer-term uptake of DATs.

**Results:** Of 10,313 records, 105 contributed to the synthesis. DATs included SMS, phone, 99DOTS, video-supported therapy and pillboxes. For ‘reach’, across various settings, cellphone access varied from 50-100% and 2-31% of PWTB were excluded from accessing DATs due to technology challenges. 36-100% of PWTB agreed to use a DAT. The weighted mean of DAT engagement over dose-days was 81% for SMS, 85% for phone, 61% for 99DOTS, 87% for pillbox and 82% for VST. Concerning ‘implementation’, the fidelity of DAT implementations was affected by technological issues such as cellphone coverage, DAT malfunction and provider-facing issues; including failure to initiate intensified patient management following low DAT engagement. Findings related to RE-AIM dimensions of ‘adoption’ and ‘maintenance’ were limited.

**Conclusion:** Our findings suggest that the ‘reach’ of DATs may be limited by a cascade of barriers, including limitations in cellphone accessibility and suboptimal sustained DAT engagement by PWTB. Video and pillbox DATs have higher levels of engagement. Implementation challenges included technological and provider-facing issues. Improving implementation outcomes may be important for TB DATs to achieve broader public health impact.

## Introduction

Despite a remarkable global recovery in TB notifications reporting, diagnosis and treatment following the disruptive impact of the COVID-19 pandemic, tuberculosis (TB) remains the leading cause of death from a single infectious agent accounting for nearly 1.3 million deaths annually [1]. Completion of tuberculosis treatment and treatment adherence are crucial to the success of tuberculosis care programmes broadly. In 2022, treatment success rates were 85% worldwide, though success rates vary substantially by country and for different subpopulations of people with TB [1]. For the last few decades, directly observed therapy (DOT) was promoted as a strategy for monitoring adherence to medications by people with TB to address the persistence of the TB global pandemic [2]. DOT requires patient self-management, frequent and timely monitoring, and surveillance at the health facility which places a substantial burden on people with TB and providers.

Digital adherence technologies (DATs), such as short message service (SMS) reminders, feature (non-smart) phones, video-supported therapy (VST), smart digital pillboxes and ingestion sensors, have the potential to address these concerns [3,4]. Understanding their effects on TB care has resulted in a large number of evaluations, particularly over the last 10 years, though evidence for effectiveness on treatment adherence and outcomes showed varied results and with limited generalisability [5,6]. VST evaluations, predominantly conducted in upper-middle and upper-income settings, suggest the DAT is promising, as do approaches that combine DATs with other non-digital approaches. DATs evaluated in low and lower-middle-income settings which focussed on pillboxes, SMS reminders and medication sleeves with phone calls (branded as “99DOTS”), show no tangible benefits on treatment outcomes. Implementation of DATs in a range of settings, including the coverage of such technologies and fidelity to the planned interventions from the provider’s perspective, is important for understanding the successes and challenges in delivering these new approaches for TB care, and particularly important given the mixed effectiveness results. Evidence synthesis for DAT implementation has not been conducted to date. The RE-AIM framework, based on five dimensions, reach, effectiveness, adoption, implementation, and maintenance, is used to characterise the public health impact of an intervention [7].

Using the RE-AIM framework, we conducted a scoping review and meta-analysis of the implementation of DATs for TB and *M. tuberculosis* infection treatment in high-income, and low-and-middle-income countries. The focus of our scoping review was on the quantitative indicators used within this framework, assessing reported implementation outcomes. A companion review covers contextual factors that influence DAT implementation [8].

## Methods

### PICOS framework and scoping review design

We followed a rigorous systematic methodology, having originally designed this study as a systematic review, then later modified it to a scoping review due to high heterogeneity in the studies and measures of interest. We adhered to the Preferred Reporting Items for Systematic Reviews and Meta-analyses Extension for Scoping Reviews (PRISMA-ScR) guidelines (supplementary appendix 1). [9]. Our published review protocol is available from PROSPERO [CRD4022326968]. Assessment of the risk of bias for the included studies has not been conducted. We used the Population, Intervention, Comparison, Outcomes, and Study (PICOS) framework to define our study population, intervention of interest, comparator (supplementary appendix 2; Table S1) and study design and used the RE-AIM framework to identify and analyse the implementation-related outcomes from studies using DATs. The study populations included people with TB, people with *M. tuberculosis* infection and other individuals such as healthcare workers, programs managers, or policymakers (subsequently referred to as healthcare providers [HCP]) who are involved in the implementation of DATs within health systems for TB care. Interventions of interest were DATs which can aid the monitoring and support of TB medication adherence. These technologies included SMS, feature phone and smartphone technologies, digital pillboxes, VST, web-based approaches and ingestion sensors. Included studies may or may not have had a comparator group. We included observational and randomised studies in this review (Table S1).

### Search Strategy

We systematically searched Medline/Ovid, Embase, Cochrane Central Register of Controlled Trials, CINAHL, Web of Science, clinicaltrials.gov, and Europe PMC, using keywords for “tuberculosis” (including both active TB disease and *M. tuberculosis* infection) and “digital adherence technology” (Table S2). A Health Librarian conducted the search. Our initial search was from January 1^st^ 2000 to April 1^st^, 2022, and was then updated to cover the period from 2^nd^ April 2022 to 25^th^ April 2023. No language restrictions were applied. Following an initial attempt, conference abstracts and grey literature, though listed in our protocol as potential data sources, were not retrieved as the quality of the data from these sources lacked the necessary detail for our implementation indicators.

### Study Screening and Data Extraction

The search results were imported into Endnote where duplicates were removed and underwent two-level blinded screening by two authors (CC, SB, MS, CK, MZ, NF) independently in Rayyan (Rayyan, Cambridge USA). Conflicts were resolved by the senior investigators (KF, RS, KS). The first level of screening involved title and abstract screening and the second level was full-text screening against prespecified inclusion/exclusion criteria (Table S3). We were inclusive in the second screen, with reasons for exclusion being: (i) not satisfying our case definition for TB and/or DATs; (ii) wrong publication type (systematic review, study protocols, trial registrations); (iii) no primary data related to quantitative implementation outcomes; or (iv) duplicates. Reasons for exclusion were noted and summarised in the PRISMA flow chart (Figure S1).

For articles satisfying the first and second-level screens, quantitative outcome data on the dimensions of the RE-AIM framework were extracted from the full text using a standard extraction form. One reviewer extracted the data which was verified by a second reviewer, and any conflicts were resolved by discussion between the reviewers, with input from a third reviewer when necessary. The extracted data included the following: study characteristics (name of the author, publication year, country, study design, study setting, the aim of the study); participant characteristics (TB disease, *M. tuberculosis* infection, drug resistance, HIV status); DAT intervention (type of DAT, duration); and quantitative outcomes data using the RE-AIM framework, as described below. Some articles initially eligible for data extraction were subsequently excluded, following attempted data extraction, as no RE-AIM quantitative outcomes data were reported. To confirm the validity of our search and identify any outstanding references, we extracted references from all systematic reviews identified from our initial search (see exclusion criteria), and these references were screened for eligibility. For one article, initially included as a preprint, we abstracted data from the supplement of the subsequently published article [10].

### RE-AIM framework

The domains of the RE-AIM framework are ‘reach’, ‘effectiveness’, ‘adoption’, ‘implementation’, and ‘maintenance’. We defined reach, among people taking treatment for TB disease or infection, as measures of engagement with a DAT into three categories: A – the proportion of people with the potential to use or receive DATs (including requirements for cellphone access); B - the proportion of people with any uptake of or engagement with DATs; C-the quality and/or duration of engagement with DATs. Adoption refers to measures of DAT engagement by HCPs. Implementation was measured through the fidelity of the intervention, that is, the extent to which the DAT intervention is operated as intended. Implementation outcomes were grouped into technology-related (the extent to which the DAT functioned as intended) and provider fidelity-related (the extent to which HCPs delivering the intervention adhered to the planned/designed intervention). Finally, the extent to which DATs become integrated into routine care was captured under maintenance (Table S4). The effectiveness domain was covered in a related review [6].

Though not originally planned, we also extracted as part of reach category A data on cellphone sharing, defined as shared vs own phone not shared, by people on treatment for TB disease or infection or potentially eligible for TB preventive therapy (e.g., household contacts of a person diagnosed with TB).

### Analysis

DAT interventions which included more than one component, for example, digital pillboxes with SMS reminders, were analysed based on the DAT considered as primary to the intervention. Binary outcomes, reflecting a consistent measure of reach indicators A and B, were summarised as the number of individuals satisfying the indicator divided by the relevant denominator, alongside a 95% confidence interval and displayed using forest plots. Where relevant, plots were stratified by income level of the country where the study was conducted, grouped as lower- or lower-middle-(L/LMIC), upper-middle-(UMIC), or high-income (HIC); DAT type; or TB type. Confidence intervals for proportions were exact Clopper-Pearson intervals, with a continuity correction of 0.5 applied for measures with zero cells.

For reach C, two main measures were extracted to capture DAT engagement over the entire treatment period: a binary measure of the proportion of individuals with DAT engagement greater than a given threshold as reported by the original studies (i.e., a cut-off point; for example, >80%, >90%, etc.) and a measure of dose-days with DAT-engagement. When binary measures were reported as less than a cut-off, the data were converted to a positive measure. A pooled estimate, by engagement level, across studies was calculated using random effects meta-analysis, across all DAT types. In sub-group analyses by DAT types, we opted not to produce summary measures due to the small number of estimates per DAT type and cut-off point. Publication bias was assessed using Doi plots in keeping with guidance for meta-analyses of proportions, using the Luis Furuya-Kanamori (LFK) index [11–13]. The second measure of dose-days with DAT-engagement was extracted from reports either as an average, often the arithmetic mean, of the measure across individuals or as an overall proportion (DAT-engagement days/all dose-days), ignoring individuals; for both measures, a standard error accounting for clustering was often not reported and could not be calculated using the available data. We therefore have summarised these data graphically using a bubble plot with the size of the bubble proportional to the number of individuals contributing to the measure. Except for studies with more than one DAT intervention group, where more than one related measure was extracted from an article, we chose a single measure that was considered most consistent with dose-days with DAT engagement. Any extracted data that did not fit into the measures described above were tabulated by income level, DAT type, and TB type.

We found limited findings on adoption and maintenance indicators; therefore, these RE-AIM dimensions were summarised narratively. For implementation, indicators were ordered by DAT type and whether it was a technology issue or related to HCP fidelity to the intervention. Technology issues were grouped, where possible, from the person with TB or HCP perspective.

## Results

### Characteristics of the included studies

Following de-duplication, 10,313 articles were identified, of which 779 satisfied the first screen and 771 were assessed for eligibility. Data extraction was attempted for 185 eligible articles, including one preprint that was not originally identified, though did fall within the database search period (Figure S1). The reference search from 68 systematic reviews yielded no articles missed in our main search. Of the 185 articles from which data extraction was attempted, 80 were excluded as no relevant data were found, leaving 105 contributing to the quantitative analysis; 90 to reach, two to adoption, 28 to implementation, and three to maintenance (Figure S1). Five studies of 105 only contributed to unplanned data summaries for cellphone ownership.

Of the 105 articles, 45, 25, and 35 were from L/LMIC, UMIC, and HIC, respectively, and 91 involved treatment for tuberculosis disease. The primary DAT types reported were VST (34), digital pillbox (22), phone (20), SMS (12), 99DOTS (7), web-based app (5) and ingestion sensor (2). In addition, three articles reported on more than one DAT type, with each DAT type used by a subset of the cohort receiving TB treatment; digital pillbox and SMS, digital pillbox and 99DOTS, and VST, digital pillbox and 99DOTS [14–16]. Findings from VST studies mostly came from HIC settings while SMS, pillbox and phone-based technologies were largely used in L/LMICs and UMICs. 99DOTS was predominantly used in L/LMIC settings and digital pillboxes were used in the widest range of country income settings. See Table 1 and Table S10.

**Table 1.** Summary of included reports by DAT type, RE-AIM indicators, country income level, and calendar year (n=105)

|  | HIC |  |  |  | UMIC |  |  |  | L/LMIC |  |  |  | Overall |  |
| --- | --- | --- | --- | --- | --- | --- | --- | --- | --- | --- | --- | --- | --- | --- |
|  | 2000-2017 |  | 2018-2023 |  | 2000-2017 |  | 2018-2023 |  | 2000-2017 |  | 2018-2023 |  | n | RE-AIM |
|  | n | RE-AIM | n | RE-AIM | n | RE-AIM | n | RE-AIM | n | RE-AIM | n | RE-AIM |  |  |
| Total number | 15 | [R; A; I] | 20 | [R; I; M] | 5 | [R; I] | 20 | [R; I; M] | 12 | [R; I] | 33 | [R; A; I] | 105 | [R; A; I; M] |
| SMS | - |  | 1 | [R] | 3 | [R; I] | 2 | [R] | 3 | [R; I] | 3 | [R; I] | 12 | [R; I] |
| Phone | 1 | [R] | - |  | - |  | 5 | [R] | 3 | [R; I] | 11 | [R; I] | 20 | [R; I] |
| 99DOTS | - |  | - |  | - |  | - |  | - |  | 7 | [R; I] | 7 | [R; I] |
| Pillbox | 4 | [R; I] | 2 | [R] | - |  | 9 | [R; I; M] | 3 | [R; I] | 4 | [R] | 22 | [R; I; M] |
| VST | 9 | [R; A; I] | 15 | [R; I; M] | 1 | [R] | 3 | [R; I] | 2 | [R; I] | 4 | [R; I] | 34 | [R; A; I; M] |
| Ingestion sensor | 1 | [I] | 1 | [R] | - |  | - |  | - |  | - |  | 2 | [R] |
| Web-based Apps | - |  | 1 | [R, A] | - |  | 1 | [M] | 1 | [R] | 2 | [R] | 5 | [R; A; M] |
| >1 DAT intervention<sup>1</sup> | - |  | - |  | 1 | [R; I] | - |  | - |  | 2 | [R; I] | 3 | [R; I] |

n represents the number of reports; HIC high-income country; UMIC upper middle-income country; L/LMIC low/low middle-income country; DAT digital adherence technology; SMS short message service; VST video-supported therapy; R Reach; A Adoption; I Implementation, M Maintenance; <sup>1</sup> these reports looked at more than one DAT type, with each DAT type used by a subset of the cohort receiving TB treatment (digital pillbox, SMS, or both [16]; digital pillbox or 99DOTS [14]; and VST, digital pillbox or 99DOTS [15] ). Note: Wei et al (2023) listed under pillbox in this table. “99DOTS” is a brand name for a DAT which uses medication sleeves with phone calls by the person with TB to document treatment adherence. Five studies only contributed to the unplanned data summaries for cellphone ownership.

### Reach A—Cellphone access among people with TB

We found that across a range of settings, access to cell phones varied from 50% (15/30) to 100% (574/574). Smartphone access accounted for some of the lower proportions of access in studies predominantly from HIC settings (Figure 1a). Between 2% (1/54) to 31% (277/891) of people with TB were excluded from studies due to technology challenges such as the inability to regularly send SMS and lack of telephone signal or internet. This was more common in L/LMIC than in HIC settings (Figure 1b). Five studies, three of which were from UMIC, reported the percentage of people with TB not eligible for using a smart pillbox due to communication impairment or being too ill (Table S5). This proportion ranged from 2% (18/891) - 9.5% (21/221) [16–20]. From the additional unplanned data collection, we found that shared phone access ranged from 4% (5/122) to 50% (98/197) in L/LMIC (8 studies) and 13% (19/145) - 39% (49/125) in UMIC (3 studies). See Figure S2.

**Figure 1:**
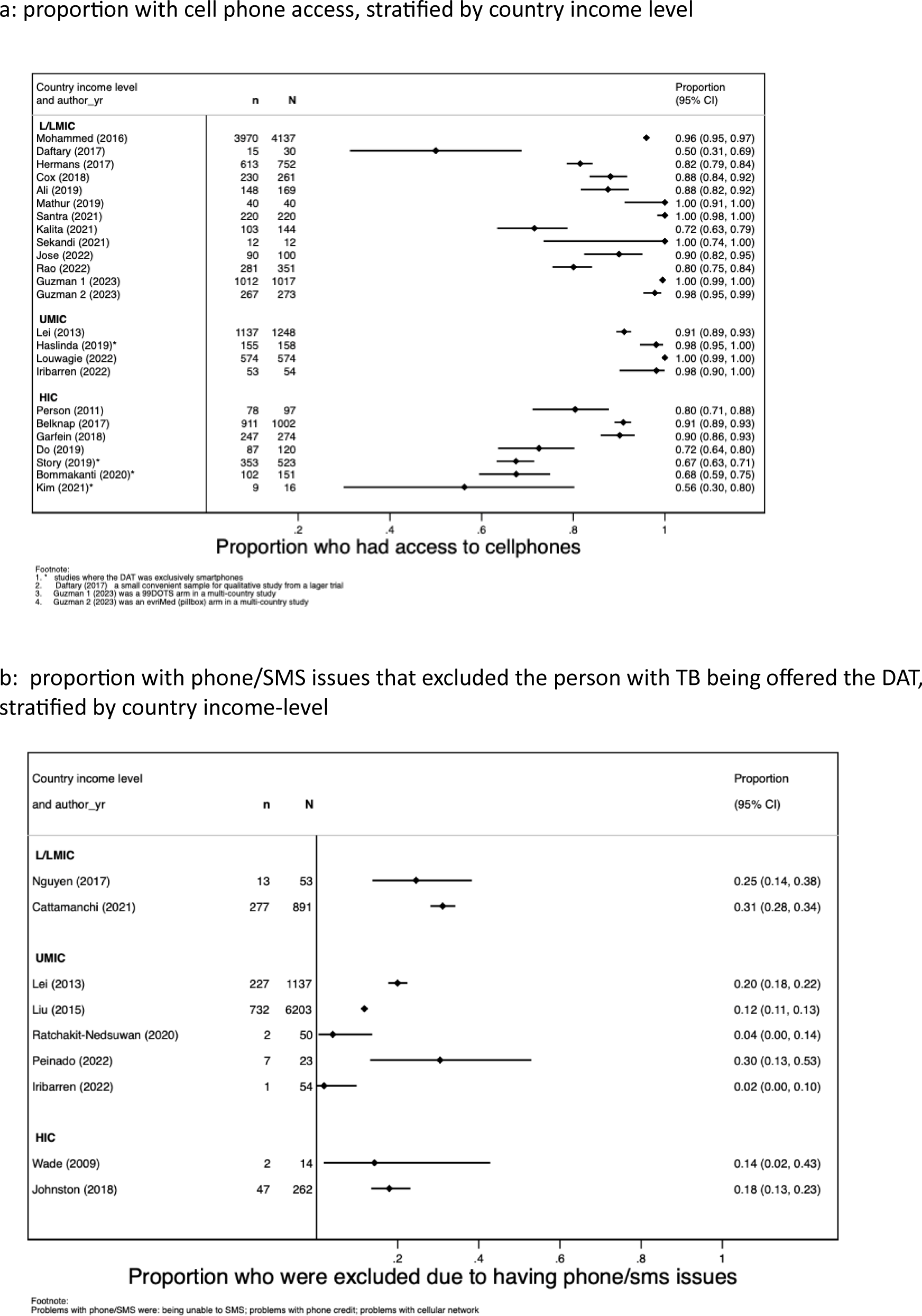
measures of Reach A – proportion with TB or *M.tuberculosis* infection, eligible to use the DAT.

TB tuberculosis, LTBI latent tuberculosis infection; DAT digital adherence technology; SMS short message service; L/LMIC low/low middle-income country; UMIC upper middle-income country; HIC high-income country.

### Reach B—Any DAT engagement by people with TB

Eight studies reported the proportion of people on TB disease treatment (plus one study for *M. tuberculosis* infection treatment) who agreed to use the DAT (Figure 2a). The percentages ranged from 36% (110/308) [21] to 100% (15/15), with the vast proportion being greater than 85%; percentages were similar by DAT type. The low uptake for VST in the study by Casalme and colleagues, conducted in Thailand, was hypothesized to have been due to the reluctance of people with TB and providers to try out new technologies [21]. Of those offered a DAT, the percentages who used the DAT at least once ranged from 33% (2746/8322) for 99DOTS to 95% (70/74) for phone-based DATs, including responses by people with TB and family members (Figure 2b). Early engagement with a DAT was measured during the first 3 months of treatment (range range 1-90 days). Measures ranged from 60% (in a 99DOTS intervention where a call was made on day 84) [22] to 89% (in a pillbox intervention were-engagement over the first 90 days for M. tuberculosis infection treatment was monitored [23]). Other varied measures of reach B are summarised in table S6.

**Figure 2:**
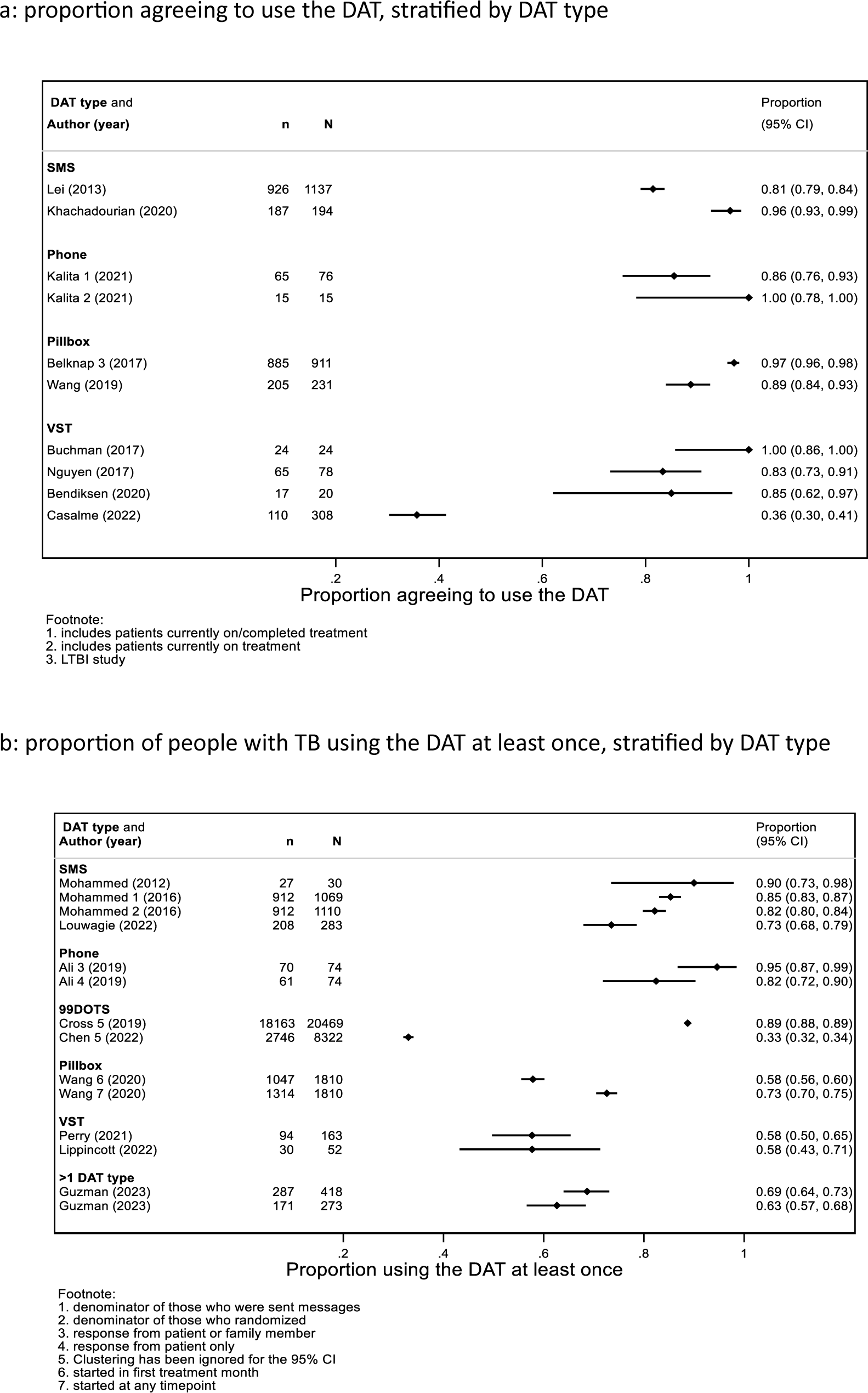
measures of Reach B – proportion of people with TB or *M.tuberculosis* infection eligible to use the DAT, who exhibit any engagement.

TB tuberculosis, DAT digital adherence technology; SMS short message service; VST video-supported therapy, 99DOTS

Four studies reported on the percentage of people with TB who stopped using the DAT during the treatment period for reasons other than death or loss to follow-up; for SMS and/or pillbox interventions the percentages ranged from 1% (12/1020) to 9.3% (9/208) [17,24,25] (Figure S3). For a VST study in the Philippines, 39.1% (43/110) stopped the DAT before they ended treatment, with 26/43 due to HCP and/or participant’s decision [21].

### Reach C—Sustained DAT engagement by people with TB

Several studies reported engagement as a proportion of individuals who achieve a specified level of engagement with the DAT. A total of 31 observations (17 studies) were included in the meta-analysis of the proportion of DAT engagement, by binary level of engagement (Figure S4). At a threshold of 100% engagement, the summary measure for the percentage of participants exhibiting perfect engagement was 29% (95%CI: 21%; 38%). The proportion of participants engaging with the DATs increased as the cut-off point reduced: at >95% cut-off, 73% (95%CI: 61%; 85%) of participants showed sustained engagement with DATs; at >90% cut-off, 85% (95% Cl: 76%; 95%) of participants; at >85% cut-off, 85% (95% Cl: 77%;94%) of participants; and at >80% cut-off, 88% (95%Cl: 75%;100%) of participants. In the subgroup analysis by DAT type, we found similarly that the proportion of individuals who achieved the level of engagement increased as the level decreased and that this was similar between DAT types (Figures S5-9). When assessing publication bias in the meta-analysis, the LFK index was less than +/-2 (-0.77) confirmed visually on the Doi plot, suggesting that effect estimates were fairly symmetrical, providing limited evidence of publication bias (Figure S11).

Other studies reported the percentage DAT engagement measured over total dose-days, either as the number of days with DAT engagement divided by the total days of observation ignoring the individual-level or an average across individuals (Figure 3, data by DAT type and income setting). The weighted average of percentage DAT engagement was 60.7% (n=6709) for 99DOTS, 80.9% (n=1201) for SMS, 81.6% (n=2179) for VST, 84.9% (n=1214) for phone and 88.3% (n=4784) for the pillbox.

**Figure 3:**
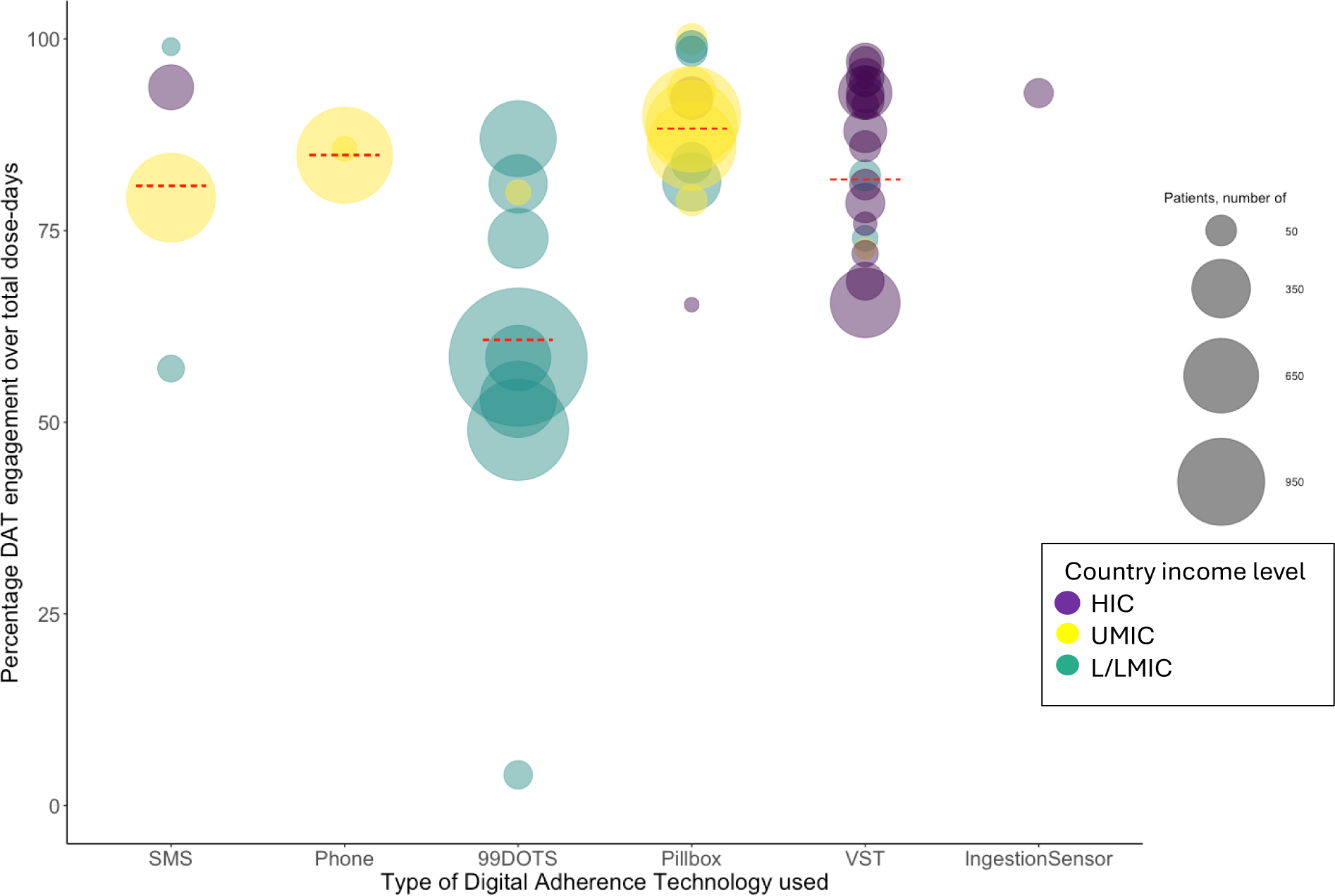
Percentage of engagement by digital adherence technology type (Reach C) [54 observations].

The plot summarises the percentage of DAT engagement over total dose days for 54 observations from 41 studies included in this analysis. Studies included for SMS interventions [16,24,26,27]; Phone [28,29]; 99DOTS [15,30–32]; Pillboxes [10,15,16,20,33–40]; VST [15,41–60]; and Ingestion Sensors [61]. For Wei (2024) and Liu (2015) pillbox openings were supplemented with pill counts so not all adherence was measured based on engagement with the digital adherence technology [10,16]. For de Groot (2022) the measure of DAT engagement as pertaining to 99DOTS and Pillbox has been adapted to remove manual dosing [15].

Country income level is indicated by the colour of the bubble and the size of the bubbles indicates the number of people with TB whose dose-days were included. The red line represents the average of percentage engagement, weighted by the sample size of the contributing study. The weighted means of DAT engagement by DAT type and country income level (number of persons with TB data included) are summarised in the table below:

|  | **SMS** | **Phone** | **99DOTs** | **Pillbox** | **VST** |
| --- | --- | --- | --- | --- | --- |
| Number of studies | 4 | 2 | 4 | 12 | 21 |
| Number of data points | 4 | 2 | 9 | 15 | 24 |
|  | Mean (n)* | Mean (n)* | Mean (n)* | Mean (n)* | Mean (n)* |
| Overall | 80.9% (1201) | 84.9% (1214) | 60.7% (6709) | 86.6% (4784) | 81.6% (2179) |
| HICs | 93.7% (170) | N/A (0) | N/A (0) | 91.7% (145) | 81.8% (1875) |
| UMICs | 79.3% (996) | 84.9% (1214) | 80.0% (22) | 88.7% (3609) | 72.6% (10) |
| L/LMICs | 63.0% (35) | N/A (0) | 60.7% (6687) | 78.8% (1030) | 80.8% (294) |

L/LMIC low/low middle-income country; UMIC upper middle-income country; HIC high-income country; SMS short message service; VST video supported therapy; \* Mean (n) represents the weighted mean (number of persons with TB).

Measures of DAT engagement that were varied or non-standard, are summarised in Table S8. These include data on adherence to a bedaquilline-based treatment regimen using pillboxes and a cut-off of >90% and >85% [20,62].

### Adoption

We found few studies that captured adoption quantitatively. Himachal Pradesh state in northern India launched its 99DOTS program in 74 TB units between 2018-9. The percentage of people with TB in each TB unit who were initiated on 99DOTS by HCPs, was measured by a proxy, which was people with TB calling 99DOTs at least once. This varied from 0.0% to 97.5% [30]. This variation in 99DOTS uptake could be interpreted as a measure of HCP adoption of the technology, assessed at the health centre level. A second study conducted in South Australia reported on the roll-out of a videophone approach for direct treatment observation from 2006 (0% [0/74] of people with TB used video service) to 2010, where 52% (30/58) of people with TB used video service [63].

### Implementation

For the implementation RE-AIM dimension, several findings were reported related to technology fidelity, the extent to which technology functioned as intended, and provider fidelity, the extent to which HCPs executed the intervention as designed/planned. Technological challenges were broadly categorised into those encountered by people with TB (e.g., phone malfunctions, flat batteries or accessories, malfunction of Apps and software, inability to use the DAT), connectivity issues (e.g., slow internet, network failure, or transmission failure) or those encountered by providers (such as poor video quality, inability to access adherence monitoring software or app) (Table 2 and Table S9).

**Table 2:**
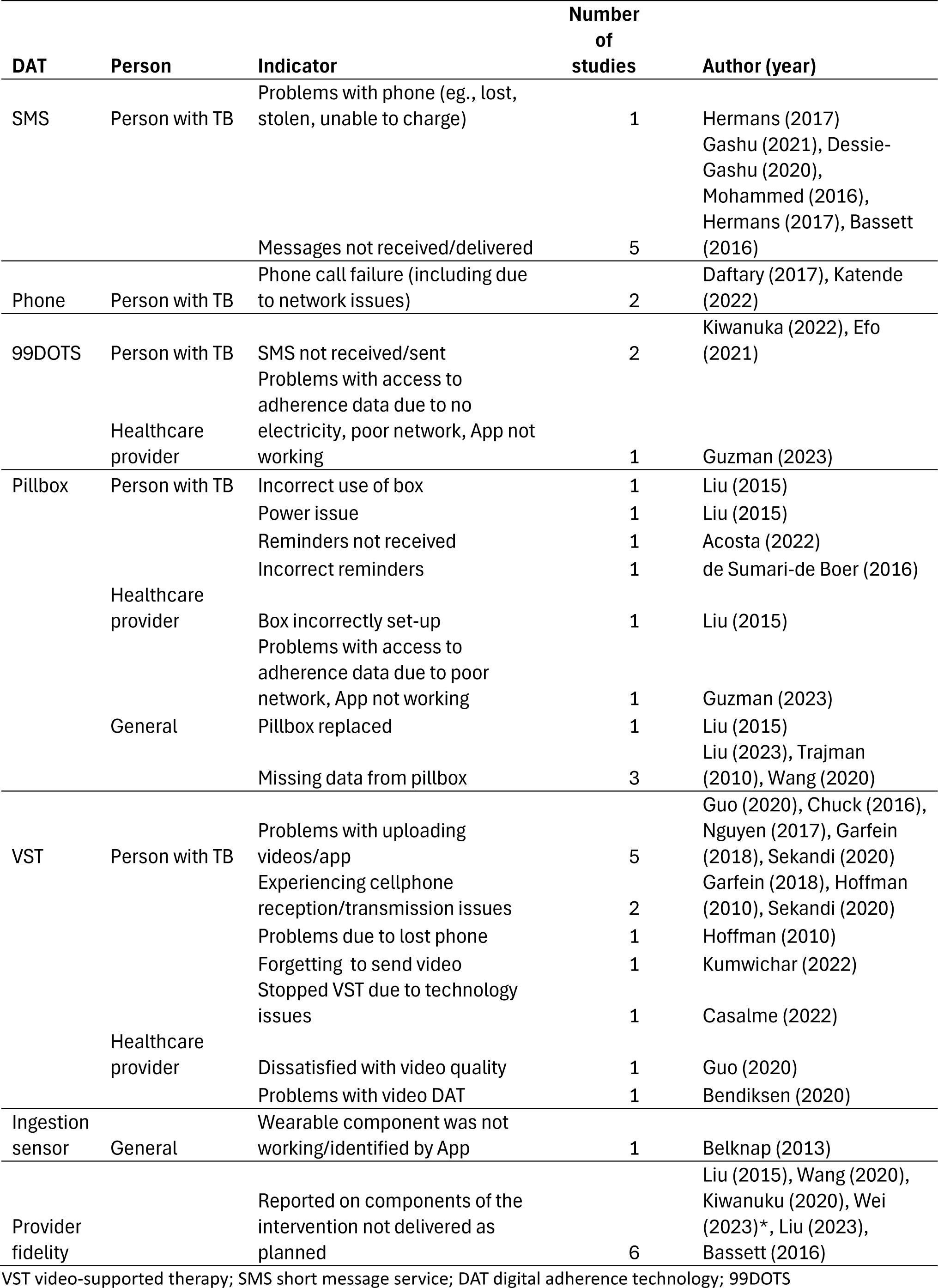
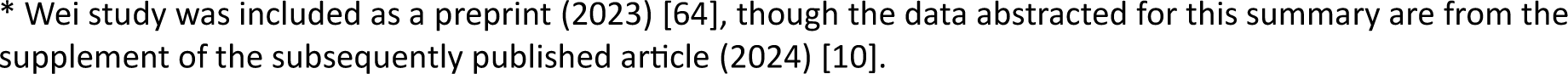
Summary of implementation indicators by DAT type.

Problems with implementing SMS-based interventions were reported by five authors [65–69]. Issues reported by people with TB included the inability to charge the phone battery, damaged phones, and no longer being able to access a shared phone. Problems with automated SMS delivery or receipt were also described. Two articles reported on technology measures with the 99DOTS intervention [14,70]. The percentage of SMSs sent among eligible days was 90%, and the percentage of SMSs received on handsets was very high (>99%). In a large multi-country study, 59% (41/70) of HCPs had problems accessing the adherence data (on a digital platform) at some point during the study period, due to lack of electricity, poor network, or the app not working, though a high percentage could access the adherence data daily (92%; 56/61). Seven studies reported implementation challenges with the pillbox [14,16,26,33,36,71,72]. Issues included incorrect pillbox set-up, lack of power, pillbox failure, and incomplete pillbox data. Similar problems with HCPs accessing adherence were reported, as for 99DOTS. Challenges with implementing VST were reported by nine authors, the majority from HIC settings [21,43,44,47,58,60,73–75]. Technology problems experienced by people with TB primarily involved VST platform/application difficulties, including uploading of videos. The percentage of people with TB experiencing this at least once ranged from 40-89%, though frequent problems were rare (8-11%). Reasons for videos not being transmitted included phone malfunction, phone loss, battery and App issues, and lack of or slow internet.

Measures of provider fidelity reflected adherence to the planned intervention by HCPs delivering the intervention (Table S9). The data extracted mainly focused on studies where the intervention included a protocolised change in management, such as a switch of people with TB to more intensive supervision and/or in-person DOT, following a display of low DAT engagement. Three studies from China that implemented a change to management reported on such issues [16,36,72]. The percentage of participants who started intensive management (regular visits by village/town doctor), of those eligible, ranged from 15.1%-82.1% (4 study arms; [16,36]). Despite the reported percentage being high in the latter study, this was not corroborated by participant self-report of change in the average number of visits per month, after intensified management. The percentage of participants who started DOT (daily observed treatment by a health practitioner), of those eligible, ranged from 3.8% (9/240) to 53.5% (53/99) [16,36,72]. Wei et al reported high levels of fidelity (>88% per month), for the change to VST among those who missed >3 consecutive doses, or for whom a doctor had concerns about their adherence [10]. Bassett et al reported fidelity of 72% (694 out of 976 participants) for the receipt ≥5 call attempts [76].

### Maintenance

An assessment of TB patient care and support policies in high-burden countries was conducted in 2018, with responses from 23 of 30 high-TB-burden countries. Digital interventions to support treatment adherence which were incorporated into the National TB Program policies included VST (3/23), pillboxes (2/23), 99DOTS (4/23), two-way SMS (2/23), and one-way SMS (2/23). Few countries, however, reported using these technologies, and often only in small-scale pilot studies [77]. A survey conducted in 2015 of US states, large cities, and US-affiliated Pacific Islands indicated that 42% (47/113) were using VST at that time, while 36% (41/113) had plans to do so [78]. A phased implementation of pillboxes for people with DS-TB adults was conducted across three provinces (138 counties) in China, beginning in 2017. By January 2019, all counties in those provinces were implementing pillboxes [79].

## Discussion

In this scoping review, we summarise quantitative findings of implementation outcomes of TB DATs using the RE-AIM framework, with most findings about dimensions of reach and implementation. Our findings provide insights into implementation challenges that might influence the effectiveness, accuracy, and real-world public health impact of these technologies. Concerning reach, successful DAT engagement by people with TB requires that multiple conditions must be fulfilled, including adequate and consistent access to cellphones or DATs, eligibility to receive a DAT, early DAT engagement, and sustained DAT engagement. We found at least moderate challenges with, or high variability in, DAT or cellphone accessibility and early and sustained DAT engagement. Together, challenges across these various levels of DAT accessibility or engagement likely result in a “cascade” of problems that may sometimes considerably limit the reach of DATs across the entire population of people being treated for TB, as shown in studies that measured reach across multiple of these levels [18,80]. Limitations in the reach of DATs may partly explain the variable effectiveness of DATs (as engagement with DATs may be important to improve adherence behaviour) and the variable accuracy of DATs (as non-engagement with DATs may result in non-reporting of doses taken) in different settings [6,81].

Our review also quantifies factors that inhibit reach such as lack of phone ownership, shared phone ownership, communication impairment (severe illness, hearing problems), lack of telephone signal or internet or “phone credit”. Secondly, relating to early and sustained engagement, the evidence suggests there is a high willingness to engage with DATs initially, but the sustained engagement seems to gradually reduce. It is possible that factors such as access (e.g., shared phones, unstable cellular network, internet or electricity supply), equity challenges (e.g., literacy, stigma) and DAT-related issues (technology fatigue, “ease of use”) could result in a reduction in engagement with the DATs. These issues are detailed in the companion review of contextual factors for DAT implementations [8]. Sub-optimal DAT function and cellular connectivity were common themes for facilitating conditions. Challenges with the use of digital technologies and connectivity have been raised as a concern for the equity of the implementation and use of DATs for TB treatment support, given that tuberculosis preferentially affects people of lower socio-economic positions [82,83]. Evidence suggested that even though cell phone coverage (Reach A) ranges from moderate to high (50%-100%), it is impacted by the ability to use a phone such as difficulty reading and sending text messages, shared ownership of phones and connectivity issues such as unreliable telephone signals or internet connection. While HIC settings recorded a slightly lower proportion with access to cellphones likely because interventions such as VST require more feature-rich mobile phones, in L/LMICs, access to cell phones may have been more affected by the connectivity issues and sharing of phones in households. On balance, we found that the proportion of people who were excluded from the use of DAT interventions was greater in LMIC than in HIC settings. We also found that individuals agreeing to use the DAT and initiating the DAT intervention (Reach B) was greatest among those using VST followed by pillboxes, lesser in SMS and 99DOTS, and lowest in phone-based DATs. For sustained engagement (Reach C), we found a similar percentage (85%) of individuals satisfying thresholds of 80, 85, and 90% of doses taken. For engagement over total dose-days, 99DOTs had the lowest level (60.7%) while pillbox had the highest (88.3%). We found little DAT engagement data by treatment phase. We opted not to use the summaries in de Groot (2022)as it was not possible to separate digital & manual dosing by type of DAT [15,15].

Compared to metrics of reach, very few studies reported quantitative measures of adoption (a measure of the interaction between technology and the HCP). However, studies that did report findings found high variability in adoption at the health centre level in the few studies reporting findings. A major reported challenge that may have affected reach or coverage was technology issues encountered by people with TB, including connectivity challenges. The inability of the DAT intervention to function as intended might have affected not only coverage but also the validation and reporting of doses and therefore, their adherence levels reported. Therefore, it is important to anticipate and address DAT-related problems (phone malfunctions, flat batteries or accessories, crashing apps, and software) by, for example, ensuring rigorous quality control and assurance in the development of devices and applications/software. An important constraint of DAT fidelity was connectivity issues, which rely on third parties such as cellular network providers to ensure a stable network. In addition, geographical obstacles may underpin challenges in connectivity on an individual level, especially in LMICs and UMICs. Concerning maintenance, even though very few studies reported on this, it seems VST, pillboxes and SMS-based interventions were the most employed. VST has been implemented mostly in Western HIC and pillboxes (with or without SMS) in Asian countries.

Implementation challenges included: technology issues experienced by people with TB (problems with phone batteries, broken/lost accessories, software malfunction, and DAT malfunction such as pillbox failure); connectivity issues (problems with transmission of videos due to unstable internet or lack of cell phone reception); and technology issues experienced by HCPs (inability to access adherence data, internet access). Provider fidelity mainly focussed on the completeness of the components of the intervention, such as failure to initiate intensified PWTB management following low DAT engagement. Overall though we found that many studies did not report on measures of implementation or fidelity to the intervention. Most reports included were research studies and few looked at the routine roll-out of DATs, and therefore maintenance indicators were scarce.

This study and the companion review on contextual factors are the first, to our knowledge, to systematically report on the implementation of DATs for the treatment of TB and *M. tuberculosis* infection. We found one protocol paper aiming to conduct a systematic review of barriers and facilitators of implementing pillboxes for TB treatment; results have not yet been reported [84]. Systematic reviews of the implementation of mHealth technologies using the RE-AIM framework, however, have been conducted in other disease areas. In an extension to a systematic review of the effectiveness of digital-supported programmes for heart disease, authors reported on RE-AIM indicators from 36 publications (27 studies) [85]. They found few studies reported on maintenance and information on intervention fidelity. A systematic review of studies assessing mobile phone-based interventions for diabetes self-management found limited data on adoption, implementation, and maintenance [86]. Strengths of our study include it being a scoping review that has been conducted to a high standard, using the same methods as those used for systematic reviews, namely the systematic identification and screening, and double data extraction from studies included in the review [87].

Limitations of this review include the relatively limited data available for some dimensions of the RE-AIM framework, suggesting that these are areas for which more research is needed in the future. A total 105 articles contributed to the quantitative synthesis of DAT implementation, using the RE-AIM framework. Most data were focused on reach (90/105), followed by implementation (28/105); few studies reported data on adoption and maintenance indicators. We were not able to assess the quality of the studies as the included reports were often not aiming to measure our outcomes of interest, though these indicators were reported as additional data from their evaluations. Other limitations relate to the disparate measures used to measure DAT engagement, complicating comparability. For the meta-analysis we have calculated the confidence intervals using exact Clopper-Pearson intervals; this will have likely resulted in narrower confidence intervals for some studies as clustering, such as individuals within health facilities, has been ignored. Given our interest was in measures commonly not presented as main outcomes, we decided not to include data from published conference abstracts as the data quality was anticipated to be of insufficient detail for our review, after an initial attempt. We similarly did not assess the grey literature which may have contributed some information to the maintenance dimension of the framework, data of which were lacking. We found no evidence of publication bias when restricting to similar measures (>90% engagement) suggesting that our estimates may be representative of engagement in practice. A strength of our study is that we assessed data from a range of studies implemented over a period when mobile phone use and connectivity have increased substantially globally. This study adds to the evidence base for DAT implementation, highlighting some of the nuances between countries and measures that are important to consider in planning implementation and designing new interventions.

## Conclusion

In conclusion, this synthesis has highlighted reach and intervention fidelity challenges to DAT implementations across varied country income settings and DAT types. These issues could inform how DAT interventions could be adapted to increase equitable access to such technologies and improve treatment outcomes.

## Contributors

RS, CC, KF, and KS conceptualized the study and designed the protocol for the scoping review. GG contributed to the design of the database search, advised on study conduct and conducted the database search. MS, SB, CC, CK, MZ, and NF led the identification of eligible articles for the review, and RS, KF, and KS resolved conflicts during the article identification process. SB, CC, KF, RS, and NF extracted relevant findings from articles included in the review. KF, CC and NF organized findings, conducted analyses and produced summaries. BP helped with organization and reporting of findings in the supplementary appendices. CC, NF and KF wrote the first draft of the manuscript. All authors provided critical revisions to the initial manuscript draft and approved the final paper.

## Data Availability

The article is a scoping review - all reported data are publicly available.

