## Supplement for "Implementation outcomes of tuberculosis digital adherence technologies: a scoping review using the RE-AIM framework"

### Supplementary material for Chilala et al. Implementation of digital adherence technologies for tuberculosis treatment adherence: a systematic review and meta-analysis.

### 1. Online supplement Appendix 1

#### 1.1. Preferred Reporting Items for Systematic reviews and Meta-Analyses extension (PRISMA) abstract checklist

| Section and Topic | Item # | Checklist item | Reported (Yes/No) |
|---|---|---|---|
| **TITLE** | | | |
| Title | 1 | Identify the report as a scoping review. | Yes |
| **BACKGROUND** | | | |
| Objectives | 2 | Provide an explicit statement of the main objective(s) or question(s) the review addresses. | Yes, last sentence of Introduction of Abstract |
| **METHODS** | | | |
| Eligibility criteria | 3 | Specify the inclusion and exclusion criteria for the review. | Yes, Methods section of Abstract, first sentence |
| Information sources | 4 | Specify the information sources (e.g. databases, registers) used to identify studies and the date when each was last searched. | Yes, Methods section of the Abstract - the number of databases used is described (details of the databases are in the main methods) |
| Risk of bias | 5 | Specify the methods used to assess risk of bias in the included studies. | Not applicable for a scoping review |
| Synthesis of results | 6 | Specify the methods used to present and synthesise results. | Yes, use of the RE-AIM framework to organise results |
| **RESULTS** | | | |

| Section and Topic | Item # | Checklist item | Reported (Yes/No) |
|---|---|---|---|
| Included studies | 7 | Give the total number of included studies and participants and summarise relevant characteristics of studies. | Yes, Results section of Abstract, first two sentences |
| Synthesis of results | 8 | Present results for main outcomes, preferably indicating the number of included studies and participants for each. If meta-analysis was done, report the summary estimate and confidence/credible interval. If comparing groups, indicate the direction of the effect (i.e. which group is favoured). | Yes, Results section of the Abstract, sentences 3-4 |
| **DISCUSSION** | | | |
| Limitations of evidence | 9 | Provide a brief summary of the limitations of the evidence included in the review (e.g. study risk of bias, inconsistency and imprecision). | Yes, Results section of the Abstract, last sentence |
| Interpretation | 10 | Provide a general interpretation of the results and important implications. | Yes, Conclusion section of the Abstract |
| **OTHER** | | | |
| Funding | 11 | Specify the primary source of funding for the review. | Yes, this has been reported separately in the journal online system |
| Registration | 12 | Provide the register name and registration number. | Yes, this is reported in methods |

*From:* Page MJ, McKenzie JE, Bossuyt PM, Boutron I, Hoffmann TC, Mulrow CD, et al. The PRISMA 2020 statement: an updated guideline for reporting systematic reviews. BMJ 2021;372:n71. doi: 10.1136/bmj.n71.
For more information, visit: http://www.prisma-statement.org/

#### 1.2. Preferred Reporting Items for Systematic reviews and Meta-Analyses extension for Scoping Reviews (PRISMA-ScR) checklist

| SECTION | ITEM | PRISMA-ScR CHECKLIST ITEM | REPORTED ON PAGE # |
|---|---|---|---|
| **TITLE** | | | |
| Title | 1 | Identify the report as a scoping review. | Reported on the title page |
| **ABSTRACT** | | | |
| Structured summary | 2 | Provide a structured summary that includes (as applicable): background, objectives, eligibility criteria, sources of evidence, charting methods, results, and conclusions that relate to the review questions and objectives. | Reported in the Abstract. The PRISMA Abstract checklist has also been completed. |
| **INTRODUCTION** | | | |
| Rationale | 3 | Describe the rationale for the review in the context of what is already known. Explain why the review questions/objectives lend themselves to a scoping review approach. | Introduction, paragraphs 2 |
| Objectives | 4 | Provide an explicit statement of the questions and objectives being addressed with reference to their key elements (e.g., population or participants, concepts, and context) or other relevant key elements used to conceptualize the review questions and/or objectives. | Introduction, paragraph 3 |
| **METHODS** | | | |
| Protocol and registration | 5 | Indicate whether a review protocol exists; state if and where it can be accessed (e.g., a Web address); and if available, provide registration information, including the registration number. | Methods section, paragraph 1 |

| SECTION | ITEM | PRISMA-ScR CHECKLIST ITEM | REPORTED ON PAGE # |
|---|---|---|---|
| Eligibility criteria | 6 | Specify characteristics of the sources of evidence used as eligibility criteria (e.g., years considered, language, and publication status), and provide a rationale. | Methods section, paragraph 1, and Online supplementary appendix 1, table S1 (PICOS) |
| Information sources* | 7 | Describe all information sources in the search (e.g., databases with dates of coverage and contact with authors to identify additional sources), as well as the date the most recent search was executed. | Methods section, subsection on "Search strategy" |
| Search | 8 | Present the full electronic search strategy for at least 1 database, including any limits used, such that it could be repeated. | Online supplementary appendix 1, table S2 |
| Selection of sources of evidence† | 9 | State the process for selecting sources of evidence (i.e., screening and eligibility) included in the scoping review. | Methods section, subsection on "Study screening and data extraction", 1$^{st}$ paragraph; & 3 |
| Data charting process‡ | 10 | Describe the methods of charting data from the included sources of evidence (e.g., calibrated forms or forms that have been tested by the team before their use, and whether data charting was done independently or in duplicate) and any processes for obtaining and confirming data from investigators. | Methods section, subsection on "Study screening and data extraction", 2$^{nd}$ paragraph |
| Data items | 11 | List and define all variables for which data were sought and any assumptions and simplifications made. | Methods section, subsection on "Re-AIM framework"; and Online supplementary appendix 1, table S4 |
| Critical appraisal of individual sources of evidence§ | 12 | If done, provide a rationale for conducting a critical appraisal of included sources of evidence; describe the methods used and how this information was used in any data synthesis (if appropriate). | The methods describe the approach – initially planned as a systematic review, which was changed to a scoping review – justification given. |
| Synthesis of results | 13 | Describe the methods of handling and summarizing the data that were charted. | Methods section, entire subsection on "Analysis": |

| SECTION | ITEM | PRISMA-ScR CHECKLIST ITEM | REPORTED ON PAGE # |
|---|---|---|---|
| **RESULTS** | | | |
| Selection of sources of evidence | 14 | Give numbers of sources of evidence screened, assessed for eligibility, and included in the review, with reasons for exclusions at each stage, ideally using a flow diagram. | Results section, subsection on “Characteristics of the included studies,” paragraph 1, Figure S1 (PRISMA flowchart), and Table 1; and table S10 |
| Characteristics of sources of evidence | 15 | For each source of evidence, present characteristics for which data were charted and provide the citations. | Results section, subsection on “Characteristics of the included studies,” paragraph 1 and Table 1 |
| Critical appraisal within sources of evidence | 16 | If done, present data on critical appraisal of included sources of evidence (see item 12). | Not done |
| Results of individual sources of evidence | 17 | For each included source of evidence, present the relevant data that were charted that relate to the review questions and objectives. | Online supplementary tables S5-S9; & figure S2-S10, appendices 2 and 3, which provide details on findings from the sources of evidence that inform meta-themes for “reach” and “adoption” |
| Synthesis of results | 18 | Summarize and/or present the charting results as they relate to the review questions and objectives. | Figures 1a, 1b, 2a, 2b, 3<br><br>Tables 2<br><br>And in the text of the results section – under each RE-AIM heading |
| **DISCUSSION** | | | |
| Summary of evidence | 19 | Summarize the main results (including an overview of concepts, themes, and types of evidence available), link to the review questions and objectives, and consider the relevance to key groups. | Our findings for each of reach, adoption, implementation and maintenance are summarized in separate paragraphs |

| SECTION | ITEM | PRISMA-ScR CHECKLIST ITEM | REPORTED ON PAGE # |
|---|---|---|---|
| Limitations | 20 | Discuss the limitations of the scoping review process. | Paragraph 6 |
| Conclusions | 21 | Provide a general interpretation of the results with respect to the review questions and objectives, as well as potential implications and/or next steps. | Last paragraph of the discussion |
| **FUNDING** | | | |
| Funding | 22 | Describe sources of funding for the included sources of evidence, as well as sources of funding for the scoping review. Describe the role of the funders of the scoping review. | Funding section |

JBI = Joanna Briggs Institute; PRISMA-ScR = Preferred Reporting Items for Systematic reviews and Meta-Analyses extension for Scoping Reviews.
* Where *sources of evidence* (see second footnote) are compiled from, such as bibliographic databases, social media platforms, and Web sites.
† A more inclusive/heterogeneous term used to account for the different types of evidence or data sources (e.g., quantitative and/or qualitative research, expert opinion, and policy documents) that may be eligible in a scoping review as opposed to only studies. This is not to be confused with *information sources* (see first footnote).
‡ The frameworks by Arksey and O'Malley (6) and Levac and colleagues (7) and the JBI guidance (4, 5) refer to the process of data extraction in a scoping review as data charting.
§ The process of systematically examining research evidence to assess its validity, results, and relevance before using it to inform a decision. This term is used for items 12 and 19 instead of "risk of bias" (which is more applicable to systematic reviews of interventions) to include and acknowledge the various sources of evidence that may be used in a scoping review (e.g., quantitative and/or qualitative research, expert opinion, and policy document).

*From:* Tricco AC, Lillie E, Zarin W, O'Brien KK, Colquhoun H, Levac D, et al. PRISMA Extension for Scoping Reviews (PRISMAScR): Checklist and Explanation. Ann Intern Med. 2018;169:467–473. doi: 10.7326/M18-0850.

### 2. Online supplement Appendix 2

Additional methods and results to accompany the manuscript *Chilala et al. Implementation of digital adherence technologies for tuberculosis treatment adherence: a systematic review and meta-analysis.*

#### 2.1. Methods

##### 2.1.1. PICOS

*Table S1. PICOS table – used to define the study question and identify eligible studies*

| | |
|---|---|
| Population | Individuals with active TB disease<br>Individuals with latent TB infection<br>All other individuals (e.g., healthcare providers, programs managers, or policymakers) who are involved in implementation of TB digital adherence technologies (DATs) within health systems.<br>Individuals from both high-income and low- and middle-income country settings. |
| Intervention | Digital adherence technologies which can support monitoring TB medication adherence as an intervention being implemented. These technologies include feature phone and smartphone technologies, video-supported therapy, digital pillboxes, and ingestible sensors. |
| Comparator | The studies may or may not have had a control or comparison group. |
| Outcome | Reach (for patients): Number and proportion of the patients who engage with a DAT.<br>Adoption (by providers or healthcare system): Number and proportion of healthcare providers who engage with the DAT,<br>Implementation (by providers and healthcare system): Fidelity of the intervention refers to the extent to which the intervention is operated as intended.<br>Maintenance (by health system): The extent to which DATs become institutionalized as part of routine care. |
| Study design | Observational studies: These may include cohort, case-control, or cross-sectional studies, case series, etc. These studies will be particularly important for extracting data on RE-AIM outcome measures.<br>Trials: These may include randomized trials and trials with a quasi-experimental design. These studies will be particularly important for extracting data on RE-AIM outcome measures. |

##### 2.1.2. Summary of search terms

*Table S2. Summary of search terms.*

| | |
|---|---|
| Tuberculosis | exp tuberculosis/ OR TB OR<br>exp mycobacterium tuberculosis/ OR MTB OR<br>Kochs disease OR<br>Phthisis OR<br>drug resistant TB OR DR TB OR<br>multidrug resistant TB OR MDRTB OR<br>extensively drug resistant TB OR XDRTB OR XDR-TB OR<br>latent TB infection OR LTBI OR<br>Anti-TB treatment OR Antitubercular Agents/ OR<br>TB preventative treatment OR TPT |
| DATs | (digital OR electronic* or virtual*) adj2 adherence) OR<br>digital technolog* OR Digital health OR digital adherence technology OR DAT OR<br>digital pill box* OR smart pill box<br>digital medication monitor OR<br><br>tele* OR tele-intervention OR telemonitor* OR teletherapy OR telehealth or teleassist* OR telebased OR tele-based OR telemedicine OR<br>eHealth OR e health OR<br>((virtual* or electronic* OR mobile or wireless) adj2 observ*) OR mhealth OR m health OR<br><br>Wireless observed therapy OR WOT OR<br>Vmalt OR<br>99DOTS OR 99 DOTS OR<br>MEMS OR Medication event monitoring system OR<br>medical electronic monitoring OR<br>monitoring sensor* OR<br>ingestible sensor*<br>monitoring device* OR<br>electronic medication package* OR<br>electronic medication monitor OR<br>medication event reminder monitor OR MERM OR<br>Artificial intelligence OR<br><br>(mobile adj (device* or health or phone*)) OR<br>smartphone* or smart phone* OR<br>cellphone* or cell phone* OR<br>Mobile technology OR<br>Or feature phone OR<br>Cellular technology OR<br>SMS OR short messag* service* OR<br>text messag* OR<br>MMS OR multimedia messag* OR<br>reminder* OR<br>video* OR videophone OR video observed therapy OR VOT OR |

| | |
|---|---|
| | vdot OR video directly observed therapy OR video DOT OR<br>video supported therapy OR VST OR<br>Webcam* OR web cam* OR<br>web based OR health IT OR<br>health ICT |

##### 2.1.3. Complete list of search terms

<u>Complete list of search terms: April 2022</u>

MEDLINE on Ovid

Translated and exported to EndNote/RIS and documented and saved and linked

2470 records on April 14, 2022

Ovid MEDLINE(R) ALL <1946 to April 13, 2022>

1 exp tuberculosis/ or mycobacterium tuberculosis/ 223226

2 Antitubercular Agents/ 40109

3 (Tuberculosis or Kochs disease or Phthisis or TB or MTB or MDRTB or XDRTB or DRTB or LTBI or directly observed treatment short course).mp. 289886

4 1 or 2 or 3 [TB CONCEPT] 295272

5 ((antitubercular agents/ or directly observed therapy/ or medication adherence/ or patient compliance/ or "treatment adherence and compliance"/) and technology/) or mobile applications/ or internet/ or cell phone/ or smartphone/ or text messaging/ or computer, handheld/ or telemedicine/ or therapy, computer-assisted/ or medical informatics applications/ 139552

6 (((digital* or electronic* or mobile or wireless* or virtual*) adj2 (adherence or medication monitor* or medication package* or observ*)) or digital technolog* or technology-based or Digital health or eHealth or e health or mhealth or m health or SMS or reminder* or short messag* service* or text messag* or MMS or multimedia messag* or MEMS or (monitor* adj2 (electronic* or sensor* or device*)) or Webcam* or web cam* or smartphone* or smart phone* or web based or health IT or health ICT or ((cell* or mobile) adj1 (device* or health or phone* or technolog*)) or video* or cellphone* or feature phone* or vdot or vmalt or tele* or dat or wot or 99dots or 99 dots or monitoring system* or ingestible sensor* or merm or artificial intelligence or ai or vot or ((digital or smart) adj (pill box* or pillbox*)) or VST).mp. 639317

7 5 or 6 [DAT CONCEPT] 705189

8 4 and 7 [TB CONCEPT AND DAT CONCEPT] 2898

9 limit 8 to yr="2000 -Current" 2470

https://proxy.library.mcgill.ca/login?url=http://ovidsp.ovid.com/ovidweb.cgi?T=JS&NEWS=N&PAGE=main&SHAREDSEARCHID=2l1i2gOeCS9cXwmAbalMaCHkWD4prRHR1lh5QIwVBJkM7ND1UnAQ7GLqy8sarvLiP

Embase on Ovid

Translated and exported to EndNote/RIS and documented and saved and linked

4363 records on April 14, 2022

Embase <1996 to 2022 Week 14>

1 exp tuberculosis/ or mycobacterium tuberculosis/ 189354

2 tuberculostatic agent/ 29551

3 (Tuberculosis or Kochs disease or Phthisis or TB or MTB or MDRTB or XDRTB or DRTB or LTBI or directly observed treatment short course).mp. 227443

4 1 or 2 or 3 [TB CONCEPT] 238274

5 ((tuberculostatic agent/ or directly observed therapy/ or medication compliance/ or patient compliance/) and technology/) or communication technology/ or exp mobile application/ or internet/ or web-based intervention/ or exp mobile phone/ or text messaging/ or personal digital assistant/ or telemedicine/ or exp teleconsultation/ or telemonitoring/ or video consultation/ or computer-assisted therapy/ or computer-assisted drug therapy/ or medical informatics/ 234981

6 (((digital* or electronic* or mobile or wireless* or virtual*) adj2 (adherence or medication monitor* or medication package* or observ*)) or digital technolog* or technology-based or Digital health or eHealth or e health or mhealth or m health or SMS or reminder* or short messag* service* or text messag* or MMS or multimedia messag* or MEMS or (monitor* adj2 (electronic* or sensor* or device*)) or Webcam* or web cam* or smartphone* or smart phone* or web based or health IT or health ICT or ((cell* or mobile) adj1 (device* or health or phone* or technolog*)) or video* or cellphone* or feature phone* or vdot or vmalt or tele* or dat or wot or 99dots or 99 dots or monitoring system* or ingestible sensor* or merm or artificial intelligence or ai or vot or ((digital or smart) adj (pill box* or pillbox*)) or VST).mp. 760427

7 5 or 6 [DAT CONCEPT] 869295

8 4 and 7 [TB CONCEPT AND DAT CONCEPT] 4511

9 limit 8 to yr="2000 -Current" 4363

https://proxy.library.mcgill.ca/login?url=http://ovidsp.ovid.com/ovidweb.cgi?T=JS&NEWS=N&PAGE=main&SHAREDSEARCHID=4qmX18vJd91JLhFRKkCNbwQdXJ1kl6DMnZbzqSytTGbtPaLEivaR11twWrpJPqIU4

CINAHL on EBSCOhost

Translated and exported to EndNote/RIS and documented

1372 records on April 14, 2022

| # | Query | Limiters/Expanders | Last Run Via | Results |
|---|---|---|---|---|
| S9 | S4 AND S7 | Limiters - Published Date: 20000101-20221231<br>Expanders - Apply equivalent subjects<br>Search modes - Boolean/Phrase | Interface - EBSCOhost Research Databases<br>Search Screen - Advanced Search<br>Database - CINAHL Plus with Full Text | 1,372 |
| S8 | S4 AND S7 | Expanders - Apply equivalent subjects<br>Search modes - Boolean/Phrase | Interface - EBSCOhost Research Databases<br>Search Screen - Advanced Search<br>Database - CINAHL Plus with Full Text | 1,444 |
| S7 | S5 OR S6 | Expanders - Apply equivalent subjects<br>Search modes - Boolean/Phrase | Interface - EBSCOhost Research Databases<br>Search Screen - Advanced Search<br>Database - CINAHL Plus with Full Text | 388,328 |
| S6 | (((digital* OR electronic* OR mobile OR wireless* OR virtual*) N2 (adherence OR "medication monitor*" OR "medication package*" OR observ*)) OR "digital technolog*" OR technology-based OR "Digital health" OR eHealth OR "e health" OR mhealth OR "m health" OR SMS OR reminder* OR "short messag* service*" OR "text messag*" | Expanders - Apply equivalent subjects<br>Search modes - Boolean/Phrase | Interface - EBSCOhost Research Databases<br>Search Screen - Advanced Search | 325,900 |

| | | | | |
|---|---|---|---|---|
| | OR MMS OR "multimedia messag*" OR MEMS OR (monitor* N2 (electronic* OR sensor* OR device*)) OR Webcam* OR "web cam*" OR smartphone* OR "smart phone*" OR "web based" OR "health IT" OR "health ICT" OR ((cell* OR mobile) N1 (device* OR health OR phone* OR technolog*)) OR video* OR cellphone* OR "feature phone*" OR vdot OR vmalt OR tele* OR dat OR wot OR 99dots OR "99 dots" OR "monitoring system*" OR "ingestible sensor*" OR merm OR "artificial intelligence" OR ai OR vot OR ((digital OR smart) W1 ("pill box*" OR pillbox*)) OR VST) | | Database - CINAHL Plus with Full Text | |
| S5 | ( (MH "Antitubercular Agents") OR (MH "Directly Observed Therapy") OR (MH "Medication Compliance") OR (MH "Patient Compliance")) AND (MH "Technology") ) OR (MH "Wireless Communications") OR (MH "Mobile Applications") OR (MH "Internet") OR (MH "Internet-Based Intervention") OR (MH "Cellular Phone+") OR (MH "Text Messaging+") OR (MH "Instant Messaging") OR (MH "Interactive Voice Response Systems") OR (MH "Videoconferencing+") OR (MH "Computers, Hand-Held+") OR (MH "Telehealth+") OR (MH "Digital Technology") OR (MH "Therapy, Computer Assisted") OR (MH "Drug Therapy, Computer Assisted") OR (MH "Medical Informatics") | Expanders - Apply equivalent subjects Search modes - Boolean/Phrase | Interface - EBSCOhost Research Databases Search Screen - Advanced Search Database - CINAHL Plus with Full Text | 123,854 |
| S4 | S1 OR S2 OR S3 | Expanders - Apply equivalent subjects Search modes - Boolean/Phrase | Interface - EBSCOhost Research Databases Search Screen - Advanced Search Database - CINAHL Plus with Full Text | 37,720 |
| S3 | (Tuberculosis OR "Kochs disease" OR Phthisis OR TB OR MTB OR MDRTB OR XDRTB OR DRTB OR LTBI OR "directly observed treatment short course") | Expanders - Apply equivalent subjects Search modes - Boolean/Phrase | Interface - EBSCOhost Research Databases Search Screen - | 37,417 |

| | | | | |
|---|---|---|---|---|
| | | | Advanced Search Database - CINAHL Plus with Full Text | |
| S2 | (MH "Antitubercular Agents") | Expanders - Apply equivalent subjects Search modes - Boolean/Phrase | Interface - EBSCOhost Research Databases Search Screen - Advanced Search Database - CINAHL Plus with Full Text | 4,730 |
| S1 | (MH "Tuberculosis+") OR (MH "Mycobacterium Tuberculosis") | Expanders - Apply equivalent subjects Search modes - Boolean/Phrase | Interface - EBSCOhost Research Databases Search Screen - Advanced Search Database - CINAHL Plus with Full Text | 25,774 |

CENTRAL on Cochrane Library

Translated and exported to EndNote/RIS and documented

1908 records on April 14, 2022

Search Name:

Date Run: 14/04/2022 19:14:58

Comment:

ID Search Hits

#1 (Tuberculosis or "Kochs disease" or Phthisis or TB or MTB or MDRTB or XDRTB or DRTB or LTBI or "directly observed treatment short course"):ti,ab,kw 8527

#2 (((digital* or electronic* or mobile or wireless* or virtual*) NEAR/2 (adherence or medication NEXT monitor* or medication NEXT package* or observ*)) or digital NEXT technolog* or technology-based or "Digital health" or eHealth or "e health" or mhealth or "m health" or SMS or reminder* or short NEXT messag* NEXT service* or text NEXT messag* or MMS or multimedia NEXT messag* or MEMS or (monitor* NEAR/2 (electronic* or sensor* or device*)) or Webcam* or web NEXT cam* or smartphone* or smart NEXT phone* or "web based" or "health IT" or "health ICT" or ((cell* or mobile) NEXT (device* or health or phone* or technolog*)) or video* or cellphone* or feature NEXT phone* or vdot or vmalt or tele* or dat or wot or 99dots or 99 NEXT dots or monitoring NEXT system* or ingestible NEXT sensor* or merm or artificial NEXT intelligence or ai or vot or ((digital or smart) NEXT (pill box* or pillbox*)) or VST):ti,ab,kw 315245

#3 #1 AND #2 with Publication Year from 2000 to 2022, in Trials 1908

[No link provided]

SCI-EXP, CPCI-S, ESCI (Web of Science Core Collection)

Translated and exported to EndNote/RIS and documented and linked

Exact search

2527 records on April 14, 2022

TS=(Tuberculosis OR "Kochs disease" OR Phthisis OR TB OR MTB OR MDRTB OR XDRTB OR DRTB OR LTBI OR "directly observed treatment short course" )

AND

TS=(((digital* OR electronic* OR mobile OR wireless* OR virtual* ) NEAR/2 (adherence OR "medication monitor*" OR "medication package*" OR observ* )) OR "digital technolog*" OR technology-based OR "Digital health" OR eHealth OR "e health" OR mhealth OR "m health" OR SMS OR reminder* OR "short messag* service*" OR "text messag*" OR MMS OR "multimedia messag*" OR MEMS OR (monitor* NEAR/2 (electronic* OR sensor* OR device* )) OR Webcam* OR "web cam*" OR smartphone* OR "smart phone*" OR "web based" OR "health IT" OR "health ICT" OR ((cell* OR mobile ) NEAR/1 (device* OR health OR phone* OR technolog* )) OR video* OR cellphone* OR "feature phone*" OR vdot OR vmalt OR tele* OR dat OR wot OR 99dots OR "99 dots" OR "monitoring system*" OR "ingestible sensor*" OR merm OR "artificial intelligence" OR ai OR vot OR ((digital OR smart ) NEAR/0 ("pill box*" OR pillbox* )) OR VST )

2000-

https://www.webofscience.com/wos/woscc/summary/24ede919-4384-46a5-92ed-3100dc08594b-310dc79f/relevance/1

MedRxiv and other preprints via Europe PMC

Translated and exported to EndNote/RIS and documented

266 records on April 19, 2022

(TITLE:Tuberculosis OR TITLE:"Kochs disease" OR TITLE:Phthisis OR TITLE:TB OR TITLE:MTB OR TITLE:MDRTB OR TITLE:XDRTB OR TITLE:DRTB OR TITLE:LTBI OR TITLE:"directly observed treatment short course" OR TITLE:"antituberculosis" OR TITLE:"antituberculous") AND (Adher* OR "directly observed" OR Digital OR electronic OR internet OR mobile OR wireless OR virtual OR TITLE:technology OR "technology based" OR tele* OR ehealth OR "e health" OR mhealth OR "m health" OR SMS OR reminder* OR messaging OR message* OR MEMS OR web OR webcam* OR smartphone* OR "health IT" OR "health ICT" OR video* OR cellphone* OR phone* OR vdot OR vmalt OR dat OR wot OR 99dots OR "99 dots" OR "monitoring system" OR "monitoring systems" OR "ingestible sensor" OR "ingestible sensors" OR merm OR "artificial intelligence" OR AI OR VOT OR "smart pillbox" OR "smart pill box" OR VST OR "computer assisted") AND (SRC:PPR)

clinicaltrials.gov

Suggested

Condition or disease:

Tuberculosis OR Kochs disease OR Phthisis OR TB OR MTB OR MDRTB OR XDRTB OR DRTB OR LTBI OR directly observed treatment short course

Other terms:

Adherence OR directly observed OR computer OR Digital OR electronic OR internet OR mobile OR virtual OR technology OR video OR mhealth OR artificial intelligence OR AI OR cellphones OR SMS OR reminders OR monitoring OR MEMS OR DAT OR sensors

<u>Complete list of search terms: April 2023</u>

| Database | Platform | Update date | # of records (total) | Notes |
|---|---|---|---|---|
| MEDLINE | Ovid | 20230421<br><br>Note: If updating after this date, start at 20230419 for consistency | 354 of 2793 | Limited to ("20220414" or "20220415" or "20220416" or "20220417" or "20220418" or "20220419" or 2022042* or 2022043* or 202205* or 202206* or 202207* or 202208* or 202209* or 20221* or 2023*).dt,ez,da. |
| Embase | Ovid | 20230421<br><br>Note: Results limited up to 20230419 | 661 of 5020 | Limited to dc=20220414-20230419 |

| | | | | |
|---|---|---|---|---|
| CINAHL | Cochrane Library/Wiley | 20230419 | 146 of 857 | Limited to EM 20220414- OR ZD "in process" |
| CENTRAL | EBSCOhost | 20230419 | 196 of 2118 | Date added to CENTRAL trials database<br><br>Custom Range: 14/04/2022 to 19/04/2023 |
| SCI-EXPANDED, CPCI-S, ESCI | Web of Science Core Collection | 20230419 | 329 of 2863 | Limited by Index date range 2022-04-14 to 2023-04-19 |
| Preprints | Europe PMC | 20230419 | 114 of 380 | CREATION_DATE:[2022-04-19 TO 2023-04-19]<br><br>Search only includes concepts of TB and DATs |
| | | | 1,800 | TOTAL |

MEDLINE (Ovid)

Ovid MEDLINE(R) ALL <1946 to April 20, 2023>

1 exp tuberculosis/ or mycobacterium tuberculosis/ 228085

2 Antitubercular Agents/ 41252

3 (Tuberculosis or Kochs disease or Phthisis or TB or MTB or MDRTB or XDRTB or DRTB or LTBI or directly observed treatment short course).mp. 300942

4 1 or 2 or 3 [TB CONCEPT] 306368

5 ((antitubercular agents/ or directly observed therapy/ or medication adherence/ or patient compliance/ or "treatment adherence and compliance"/) and technology/) or mobile applications/ or internet/ or cell phone/ or smartphone/ or text messaging/ or computer, handheld/ or telemedicine/ or therapy, computer-assisted/ or medical informatics applications/ 148180

6 (((digital* or electronic* or mobile or wireless* or virtual*) adj2 (adherence or medication monitor* or medication package* or observ*)) or digital technolog* or technology-based or Digital health or eHealth or e health or mhealth or m health or SMS or reminder* or short messag* service* or text messag* or MMS or multimedia messag* or MEMS or (monitor* adj2 (electronic* or sensor* or device*)) or Webcam* or web cam* or smartphone* or smart phone* or web based or health IT or health ICT or ((cell* or mobile) adj1 (device* or health or phone* or technolog*)) or video* or cellphone* or feature phone* or vdot or vmalt or tele* or dat or wot or 99dots or 99 dots or monitoring system* or ingestible sensor* or merm or artificial intelligence or ai or vot or ((digital or smart) adj (pill box* or pillbox*)) or VST).mp. 705541

7 5 or 6 [DAT CONCEPT] 772972

8 4 and 7 [TB CONCEPT AND DAT CONCEPT] 3221

9 limit 8 to yr="2000 -Current" 2793

10 ("20220414" or "20220415" or "20220416" or "20220417" or "20220418" or "20220419" or 2022042* or 2022043* or 202205* or 202206* or 202207* or 202208* or 202209* or 20221* or 2023*).dt,ez,da. 1903196

11 9 and 10 354

https://proxy.library.mcgill.ca/login?url=https://ovidsp.ovid.com/ovidweb.cgi?T=JS&NEWS=N&PAGE=main&SHAREDSEARCHID=3fnvVdcJME85FE7alzUoGwgVxZWFQGclstIdKYE9CMQFMhddHUd1jxgKmo1hQDBKW

Embase (Ovid)

Embase <1996 to 2023 Week 15>

1 exp tuberculosis/ or mycobacterium tuberculosis/ 204337

2 tuberculostatic agent/ 30987

3 (Tuberculosis or Kochs disease or Phthisis or TB or MTB or MDRTB or XDRTB or DRTB or LTBI or directly observed treatment short course).mp. 246187

4 1 or 2 or 3 [TB CONCEPT] 257637

5 ((tuberculostatic agent/ or directly observed therapy/ or medication compliance/ or patient compliance/) and technology/) or communication technology/ or exp mobile application/ or internet/ or web-based intervention/ or exp mobile phone/ or text messaging/ or personal digital assistant/ or telemedicine/ or exp teleconsultation/ or telemonitoring/ or video consultation/ or computer-assisted therapy/ or computer-assisted drug therapy/ or medical informatics/ 261570

6 (((digital* or electronic* or mobile or wireless* or virtual*) adj2 (adherence or medication monitor* or medication package* or observ*)) or digital technolog* or technology-based or Digital health or eHealth or e health or mhealth or m health or SMS or reminder* or short messag* service* or text messag* or MMS or multimedia messag* or MEMS or (monitor* adj2 (electronic* or sensor* or device*)) or Webcam* or web cam* or smartphone* or smart phone* or web based or health IT or health ICT or ((cell* or mobile) adj1 (device* or health or phone* or technolog*)) or video* or cellphone* or feature phone* or vdot or vmalt or tele* or dat or wot or 99dots or 99 dots or monitoring system* or ingestible sensor* or merm or artificial intelligence or ai or vot or ((digital or smart) adj (pill box* or pillbox*)) or VST).mp. 870210

7 5 or 6 [DAT CONCEPT] 984663

8 4 and 7 [TB CONCEPT AND DAT CONCEPT] 5167

9 limit 8 to yr="2000 -Current" 5020

10 limit 9 to dc=20220414-20230419 661

https://proxy.library.mcgill.ca/login?url=https://ovidsp.ovid.com/ovidweb.cgi?T=JS&NEWS=N&PAGE=main&SHAREDSEARCHID=7KKR39ofVynipL3CX0ToO3GWb5GrslsDhbjFanUYdIRYp9ugDOcxhfduODOtnrlX1

CINAHL (EBSCOhost)

| # | Query | Limiters/Expanders | Last Run Via | Results |
|---|---|---|---|---|
| S11 | S9 AND S10 | Expanders - Apply equivalent subjects Search modes - Boolean/Phrase | Interface - EBSCOhost Research Databases Search Screen - Advanced Search Database - CINAHL Plus with Full Text | 146 |
| S10 | EM 20220414- OR ZD "in process" | Expanders - Apply equivalent subjects Search modes - Boolean/Phrase | Interface - EBSCOhost Research Databases Search Screen - Advanced Search Database - CINAHL Plus with Full Text | 1,147,669 |
| S9 | S4 AND S7 | Limiters - Published Date: 20000101-20231231 Expanders - Apply equivalent subjects Search modes - Boolean/Phrase | Interface - EBSCOhost Research Databases Search Screen - Advanced Search Database - CINAHL Plus with Full Text | 857 |

| | | | | |
|---|---|---|---|---|
| S8 | S4 AND S7 | Expanders - Apply equivalent subjects Search modes - Boolean/Phrase | Interface - EBSCOhost Research Databases Search Screen - Advanced Search Database - CINAHL Plus with Full Text | 916 |
| S7 | S5 OR S6 | Expanders - Apply equivalent subjects Search modes - Boolean/Phrase | Interface - EBSCOhost Research Databases Search Screen - Advanced Search Database - CINAHL Plus with Full Text | 347,115 |
| S6 | (((digital* OR electronic* OR mobile OR wireless* OR virtual*) N2 (adherence OR "medication monitor*" OR "medication package*" OR observ*)) OR "digital technolog*" OR technology-based OR "Digital health" OR eHealth OR "e health" OR mhealth OR "m health" OR SMS OR reminder* OR "short messag* service*" OR "text messag*" OR MMS OR "multimedia messag*" OR MEMS OR (monitor* N2 (electronic* OR sensor* OR device*)) OR Webcam* OR "web cam*" OR smartphone* OR "smart phone*" OR "web based" OR "health IT" OR "health ICT" OR ((cell* OR mobile) N1 (device* OR health OR phone* OR technolog*)) OR video* OR cellphone* OR "feature phone*" OR vdot OR vmalt OR tele* OR dat OR wot OR 99dots OR S99 dots OR "monitoring system*" OR "ingestible sensor*" OR merm OR "artificial intelligence" OR ai OR vot OR ((digital OR smart) W1 ("pill box*" OR pillbox*)) OR VST) | Expanders - Apply equivalent subjects Search modes - Boolean/Phrase | Interface - EBSCOhost Research Databases Search Screen - Advanced Search Database - CINAHL Plus with Full Text | 278,594 |

| | | | | |
|---|---|---|---|---|
| S5 | ( (MH "Antitubercular Agents") OR (MH "Directly Observed Therapy") OR (MH "Medication Compliance") OR (MH "Patient Compliance")) AND (MH "Technology") ) OR (MH "Wireless Communications") OR (MH "Mobile Applications") OR (MH "Internet") OR (MH "Internet-Based Intervention") OR (MH "Cellular Phone+") OR (MH "Text Messaging+") OR (MH "Instant Messaging") OR (MH "Interactive Voice Response Systems") OR (MH "Videoconferencing+") OR (MH "Computers, Hand-Held+") OR (MH "Telehealth+") OR (MH "Digital Technology") OR (MH "Therapy, Computer Assisted") OR (MH "Drug Therapy, Computer Assisted") OR (MH "Medical Informatics") | Expanders - Apply equivalent subjects Search modes - Boolean/Phrase | Interface - EBSCOhost Research Databases Search Screen - Advanced Search Database - CINAHL Plus with Full Text | 133,190 |
| S4 | S1 OR S2 OR S3 | Expanders - Apply equivalent subjects Search modes - Boolean/Phrase | Interface - EBSCOhost Research Databases Search Screen - Advanced Search Database - CINAHL Plus with Full Text | 39,412 |
| S3 | (Tuberculosis OR "Kochs disease" OR Phthisis OR TB OR MTB OR MDRTB OR XDRTB OR DRTB OR LTBI OR "directly observed treatment short course") | Expanders - Apply equivalent subjects Search modes - Boolean/Phrase | Interface - EBSCOhost Research Databases Search Screen - Advanced Search Database - CINAHL Plus with Full Text | 39,091 |
| S2 | (MH "Antitubercular Agents") | Expanders - Apply equivalent subjects | Interface - EBSCOhost Research Databases | 4,883 |

| | | | | |
|---|---|---|---|---|
| | | Search modes - Boolean/Phrase | Search Screen - Advanced Search Database - CINAHL Plus with Full Text | |
| S1 | (MH "Tuberculosis+") OR (MH "Mycobacterium Tuberculosis") | Expanders - Apply equivalent subjects Search modes - Boolean/Phrase | Interface - EBSCOhost Research Databases Search Screen - Advanced Search Database - CINAHL Plus with Full Text | 26,642 |

CENTRAL (Cochrane Library/Wiley)

Search Name:

Date Run: 19/04/2023 19:42:11

Comment:

ID Search Hits

#1 (Tuberculosis or "Kochs disease" or Phthisis or TB or MTB or MDRTB or XDRTB or DRTB or LTBI or "directly observed treatment short course"):ti,ab,kw 9141

#2 (((digital* or electronic* or mobile or wireless* or virtual*) NEAR/2 (adherence or medication NEXT monitor* or medication NEXT package* or observ*)) or digital NEXT technolog* or technology-based or "Digital health" or eHealth or "e health" or mhealth or "m health" or SMS or reminder* or short NEXT messag* NEXT service* or text NEXT messag* or MMS or multimedia NEXT messag* or MEMS or (monitor* NEAR/2 (electronic* or sensor* or device*)) or Webcam* or web NEXT cam* or smartphone* or smart NEXT phone* or "web based" or "health IT" or "health ICT" or ((cell* or mobile) NEXT (device* or health or phone* or technolog*)) or video* or cellphone* or feature NEXT phone* or vdot or vmalt or tele* or dat or wot or 99dots or 99 NEXT dots or monitoring NEXT system* or ingestible NEXT sensor* or merm or artificial NEXT intelligence or ai or vot or ((digital or smart) NEXT (pill box* or pillbox*)) or VST):ti,ab,kw 359210

#3 #1 AND #2 with Publication Year from 2000 to 2023, in Trials 2118

Date added to CENTRAL trials database

Custom Range: 14/04/2022 to 19/04/2023

196 of 2118 records

WOS.SCI,WOS.ISTP,WOS.ESCI (Web of Science Core Collection)

### Web of Science Search Strategy (v0.1)

Search: TS=(Tuberculosis OR "Kochs disease" OR Phthisis OR TB OR MTB OR MDRTB OR XDRTB OR DRTB OR LTBI OR "directly observed treatment short course" )

AND

TS=(((digital* OR electronic* OR mobile OR wireless* OR virtual* ) NEAR/2 (adherence OR "medication monitor*" OR "medication package*" OR observ* )) OR "digital technolog*" OR technology-based OR "Digital health" OR eHealth OR "e health" OR mhealth OR "m health" OR SMS OR reminder* OR "short messag* service*" OR "text messag*" OR MMS OR "multimedia messag*" OR MEMS OR (monitor* NEAR/2 (electronic* OR sensor* OR device* )) OR Webcam* OR "web cam*" OR smartphone* OR "smart phone*" OR "web based" OR "health IT" OR "health ICT" OR ((cell* OR mobile ) NEAR/1 (device* OR health OR phone* OR technolog* )) OR video* OR cellphone* OR "feature phone*" OR vdot OR vmalt OR tele* OR dat OR wot OR 99dots OR "99 dots" OR "monitoring system*" OR "ingestible sensor*" OR merm OR "artificial intelligence" OR ai OR vot OR ((digital OR smart ) NEAR/0 ("pill box*" OR pillbox* )) OR VST )

Editions: WOS.SCI,WOS.ISTP,WOS.ESCI

Timespan: 2000-01-01 to 2023-04-19

Date Run: Fri Apr 21 2023 14:19:23 GMT-0400 (Eastern Daylight Time)

Results: 2863

### Database: Web of Science Core Collection

### Entitlements:

- WOS.IC: 1993 to 2023

- WOS.CCR: 1985 to 2023

- WOS.SCI: 1900 to 2023

- WOS.AHCI: 1975 to 2023

- WOS.BHCI: 2005 to 2023

- WOS.BSCI: 2005 to 2023

- WOS.ESCI: 2005 to 2023

- WOS.ISTP: 1990 to 2023

- WOS.SSCI: 1900 to 2023

- WOS.ISSHP: 1990 to 2023

### Searches:

Search:

#1

Timespan: 2022-04-14 to 2023-04-19

Date Run: Fri Apr 21 2023 14:20:33 GMT-0400 (Eastern Daylight Time)

Results: 329

MedRxiv and other preprints via Europe PMC

(TITLE:Tuberculosis OR TITLE:"Kochs disease" OR TITLE:Phthisis OR TITLE:TB OR TITLE:MTB OR TITLE:MDRTB OR TITLE:XDRTB OR TITLE:DRTB OR TITLE:LTBI OR TITLE:"directly observed treatment short course" OR TITLE:"antituberculosis" OR TITLE:"antituberculous") AND (Adher* OR "directly observed" OR Digital OR electronic OR internet OR mobile OR wireless OR virtual OR TITLE:technology OR "technology based" OR tele* OR ehealth OR "e health" OR mhealth OR "m health" OR SMS OR reminder* OR messaging OR message* OR MEMS OR web OR webcam* OR smartphone* OR "health IT" OR "health ICT" OR video* OR cellphone* OR phone* OR vdot OR vmalt OR dat OR wot OR 99dots OR "99 dots" OR "monitoring system" OR "monitoring systems" OR "ingestible sensor" OR "ingestible sensors" OR merm OR "artificial intelligence" OR AI OR VOT OR "smart pillbox" OR "smart pill box" OR VST OR "computer assisted") AND (SRC:PPR) AND CREATION_DATE:[2022-04-19 TO 2023-04-19]

##### 2.1.4. Inclusion and exclusion criteria

*Table S3. inclusion/exclusion criteria.*

| Inclusion criteria | Exclusion Criteria |
|---|---|
| Studies that address either active TB disease or latent TB infection in humans | Studies don't involve use of digital health interventions for tuberculosis treatment support. |
| Involve use of digital adherence technologies (DATs) in research or routine implementation | If the same study with the same outcomes has been reported by different papers, the most recent published paper will be included. |
| Studies that report data on implementation of DATs relevant to at least one dimension of the RE-AIM framework<br><br>(excluding effectiveness) | Situations in which the abstract, paper, or report does not contain sufficient information to extract. |

| | |
|---|---|
| All observational studies (cohort studies, cross sectional studies, case-control studies), trials (randomized controlled<br><br>trials) | Editorials or review articles that do not present any primary research findings. |
| Relevant abstracts and grey literature | |
| High-quality data from recent initiatives such as TB REACH | |

##### 2.1.5. RE-AIM framework for the DAT review

*Table S4. RE-AIM framework.*

| RE-AIM DOMAIN | Definition | Categories | Description | Detail |
|---|---|---|---|---|
| Reach | Number and proportion of the **patients** who **engage with a DAT**, and related contextual factors | Category A – People with TB or taking LTBI with potential to use or receive DATs | Among all people with TB/ taking LTBI, the proportion (n/N) deemed eligible to receive DATs. | People who fall into the gap in category A may not be able to use DATs due to lack of phone access, severity of TB illness or hospital admission, for example. |
| | | Category B – People with TB with any uptake of, or engagement with, DATs | Among people who are eligible to receive DATs, the proportion provided DATs OR the proportion who exhibit any engagement after having been provided DATs:<br>B1: proportion agreeing to use DAT, among those eligible<br>B2: proportion engaged with DAT at least once, among those who started.<br>B3: proportion with early engagement (over a shorter time period than entire treatment length) | People who fall into the gap may have been eligible for DATs by TB program guidelines but not actually provided with DATs, OR they may have been provided with DATs but not actually ever use the DAT (e.g., never calling 99DOTS, sending back a SMS to report a dose, opening the pillbox, etc.).<br><br>We will also include, for example, measures that look at any engagement within the first one or two months. |
| | | REACH Category C – Quality and/or duration of engagement with DATs | Among people who are provided with DATs, the proportion who exhibit various levels or durations of use over the treatment course.<br><br>C1 Binary indicator (% pts with at least xx% engagement)<br>C2 Dose-level - % engagement with DAT<br>C3 % pts who stopped using DAT (excludes deaths, LTFU) | Measures may look at overall engagement measured as (i) a percentage DAT engagement over total dose-days or (ii) percentage of patients with DAT engagement >threshold.<br><br>May look at the duration over which patients engaged (i.e., based on when patients stopped calling). |

| | | | | |
|---|---|---|---|---|
| Adoption | Number and proportion of **healthcare providers** who **engage with the DAT,** and related contextual factors. | - | the proportion of providers who have the potential to engage with the DAT intervention | |
| Implementation | Fidelity of the intervention refers to the **extent to which the intervention is operated as intended**. | Implementation Technology fidelity | Ability of the technology (DAT) to function as it was intended to. | Factors relating to network coverage, battery problems, etc. |
| | | Implementation Provider fidelity | - Consistency and delivery of the healthcare providers to the intervention, as was intended. | Delivery of DAT counselling to patients, regular review of adherence data, appropriate outreach to patients, shifting of patients back to DOT/intensive management if required. |
| Maintenance | The **extent to which DATs become institutionalized as part of routine care**. | | Long term uptake and sustainment | |

#### 2.2. Results

##### 2.2.1. Additional figures and tables

*Figure S1* - PRISMA Flow diagram of how papers were selected for inclusion into the analysis.

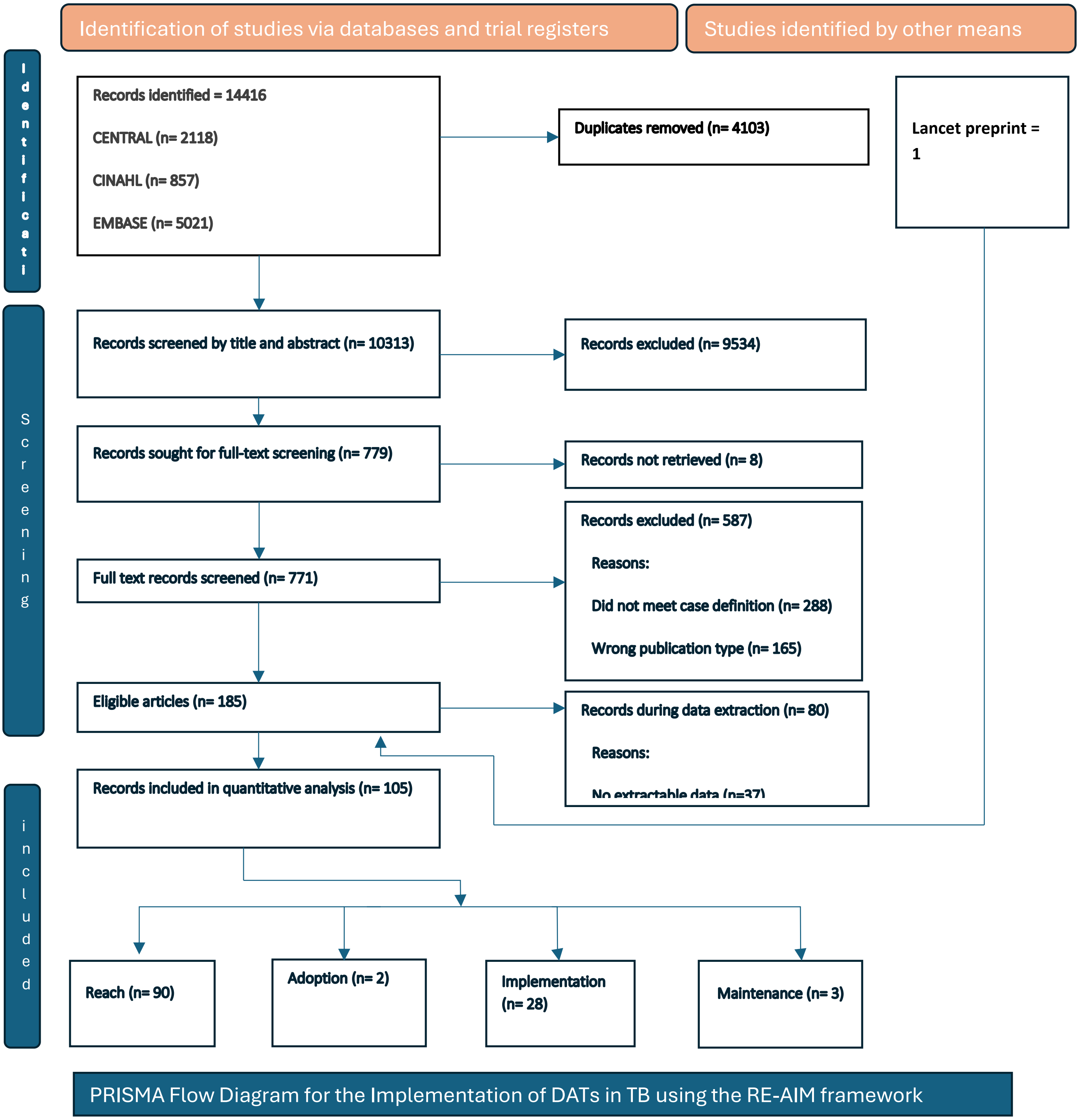

*Table S5: summary of people with TB excluded from intervention due to communication impairment (Reach A)*

| Author | Description of numerator | Description of denominator | n | N | % |
|---|---|---|---|---|---|
| Liu 2015 | Number of PWTB who were deemed unable to use SMS texting or pillbox due to communication impairment | Number of PWTB approached for the trial across all study arms | 191 | 6203 | 3% |
| Zelnick 2021 | Number of prisoners, those lacking capacity, severe hearing impairment | enrolled plus numerator | 21 | 221 | 9.5% |
| Wang 2020 | Number of PWTB not eligible due to communication impairment | all PWTB registered for TB | 162 | 2294 | 7% |
| Wang 2019 | Number of PWTB who did not start pillbox due to communication impairment | Number of PWTB who were eligible for the pillbox | 7 | 316 | 2% |
| Cattamanchi 2021 | Number of PWTB excluded due to being too sick or elderly | total number of PWTB enrolled | 18 | 891 | 2% |

PWTB person with TB; SMS short message service

*Figure S2. Proportion sharing phone, of everyone with access to a phone (unplanned data collection)*

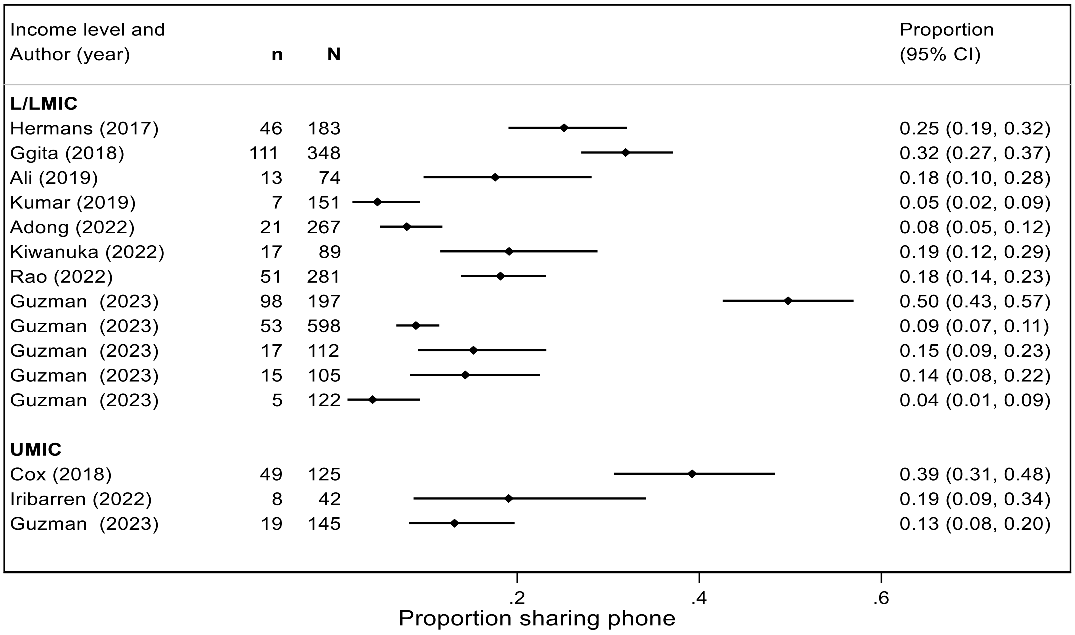

Footnote: Multiple records for Guzman reflect different countries from this multi-country study. Two studies, Musiimenta (2020) & Saunders (2018), included in the unplanned data collection did not have data on shared phone access. The proportion of people of owned phones were 89% (47/53) and 82% (2177/2655) respectively.

*Figure S3. Proportion of PWTB electing to stop using the DAT for reasons other than death or lost to follow-up*

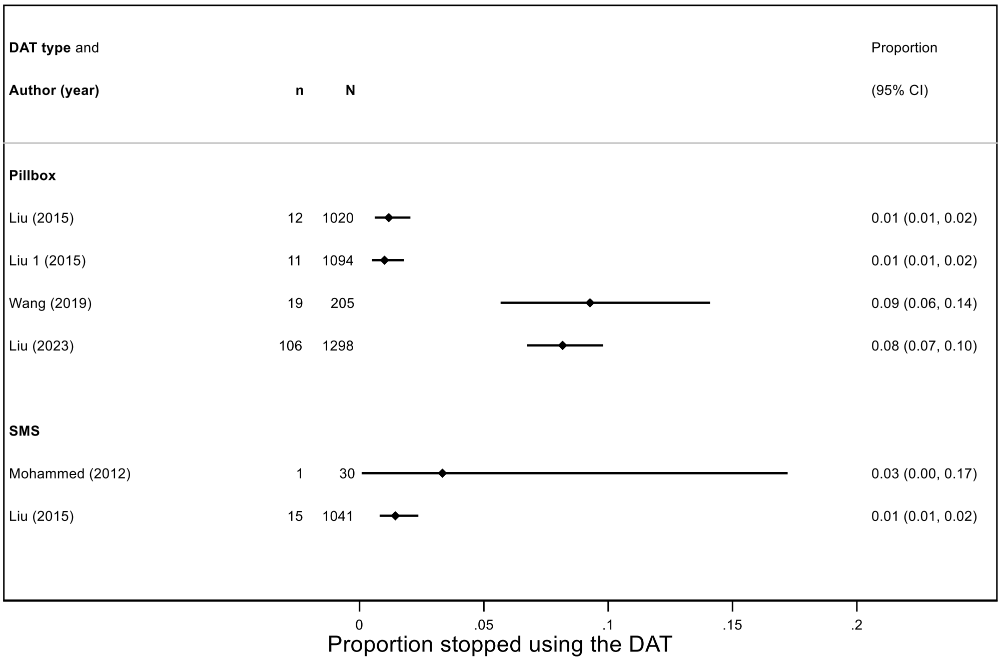

Footnote:
1. proportion stopped using the combined pillbox and SMS DAT reported under pillbox

SMS short message service
Excluded from the plot: Casalme - 43/110 (39%) stopped using vide-supported therapy (VST) prior to the treatment outcome - 26/42 moved back to DOT based on HCP/patient decision - reasons included a preference for DOT or other synchronous VST, persistent non-adherence, broken phone, change of treatment centre.

*Table S6: summary of other measures of DAT engagement (Reach B)*

| Author | Year | DAT | Setting | Patient or dose-day level [n*] | Description of Measure | Numerator | Denominator | % |
|---|---|---|---|---|---|---|---|---|
| Cross | 2019 | 99DOTS | LMIC | PWTB | % engagement on day 1 | | 20469 | 74.0% |
| Cross | 2019 | 99DOTS | LMIC | PWTB | % engagement on day 84 | | 20469 | 60.0% |
| Cross | 2019 | 99DOTS | LMIC | PWTB | % engagement on day 167 | | 20469 | 56.0% |
| Thomas | 2020 | 99DOTS | LMIC | PWTB | % engagement in the continuation phase | 252 | 394 | 64.0% |
| Thomas | 2020 | 99DOTS | LMIC | PWTB | % engagement in the intensive phase | 144 | 203 | 70.9% |
| Subbaraman | 2021 | 99DOTS | LMIC | PWTB | % with 3/3 days of non-engagement (patient-reported doses) | 191 | 608 | 31.4% |
| Subbaraman | 2021 | 99DOTS | LMIC | PWTB | % with 3/3 days of non-engagement (patient- and provider-reported doses) | 68 | 608 | 11.2% |
| Zhadnova | 2020 | phone | UMIC | PWTB | % daily DAT responses, in the first 2 months | 27 | 44 | 61.4% |
| Zhadnova | 2020 | phone | UMIC | PWTB | % responded to the DAT once every 3-7 days, in the first 2 months | 17 | 44 | 38.6% |
| Story | 2019 | VST | HIC | PWTB | % who engage with DAT for at least 1 week | 101 | 112 | 90.2% |
| Holzman | 2019 | VST | LMIC | PWTB | % who satisfied the 10-day run-in period which evaluated their ability to record and submit videos | 25 | 35 | 71.4% |
| Kiwanuka | 2022 | 99DOTS | LIC | dose-day [n=456] | median % of dose-days with engagement, during the first month | | | 71.4% |
| Kiwanuka | 2022 | 99DOTS | LIC | dose-day [n=412] | median % of dose-days with engagement, during the sixth month | | | 46.4% |

| | | | | | | | | |
|---|---|---|---|---|---|---|---|---|
| Muyindike | 2022 | pillbox | LIC | dose-day [n=275] | mean % of dose-days with engagement in the previous 90 days at 3 months | | | 89.0% |
| Muyindike | 2022 | pillbox | LIC | dose-day [n=264] | mean % of dose-days with engagement in the previous 90 days at 6 months | | | 81.0% |
| Wei | 2023 | pillbox | UMIC | PWTB | % with >80% DAT engagement in month 1 | 141 | 142 | 99.3% |
| Wei | 2023 | pillbox | UMIC | PWTB | % with >80% DAT engagement in month 2 | 140 | 142 | 98.6% |

* n represents the number of participants contributing to the dose-day measure; PWTB person with TB ; VST video-supported therapy; LIC low-income country, LMIC low-middle income country; UMIC upper-middle income country; HIC high-income country; DAT digital adherence technology

*Figure S4 Percentage of individuals with DAT engagement, by levels of engagement (Reach C)*

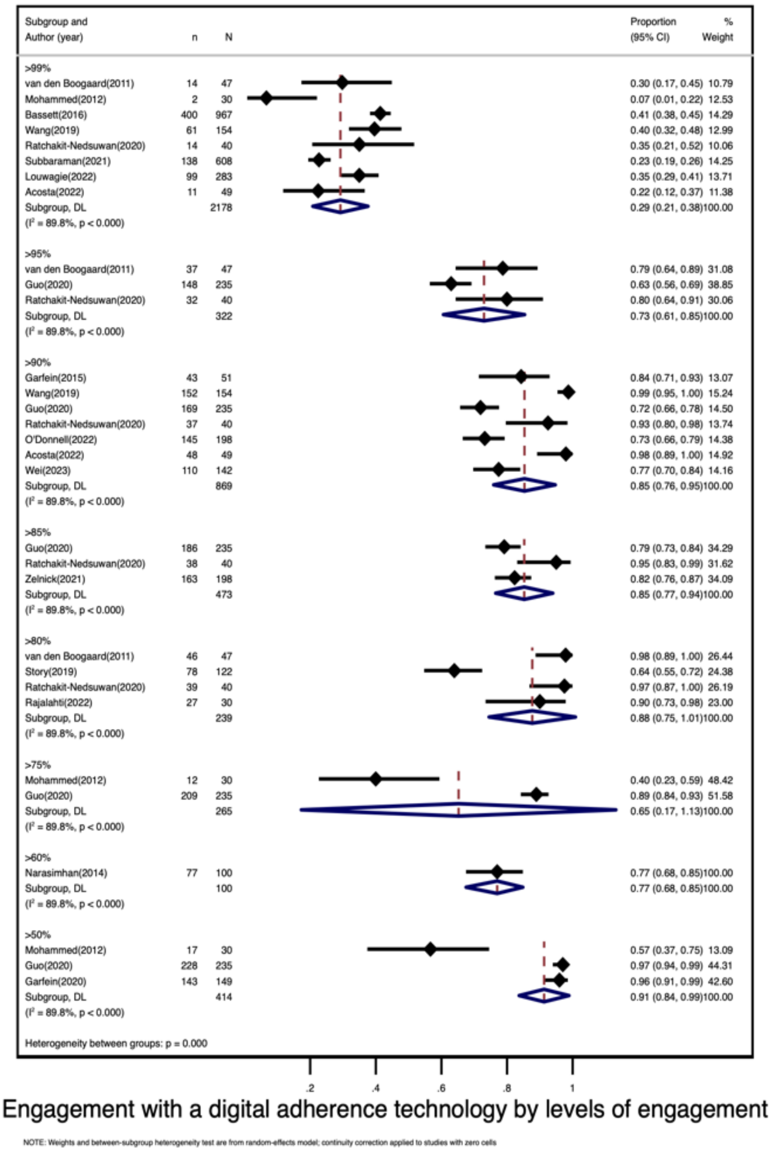

Footnote

| Level of engagement threshold | # of studies | # of persons with TB |
|---|---|---|
| >99% | 8 | 2178 |
| >95% | 3 | 322 |
| >90% | 7 | 869 |
| >85% | 3 | 473 |
| >80% | 4 | 239 |
| >75% | 2 | 265 |
| >60% | 1 | 100 |
| >50% | 3 | 414 |

**Subgroup analyses.**

Subgroup analysis, looking at levels of engagement, by DAT-type.

*Figure S5. Sustained engagement with a DAT by levels of engagement - SMS*

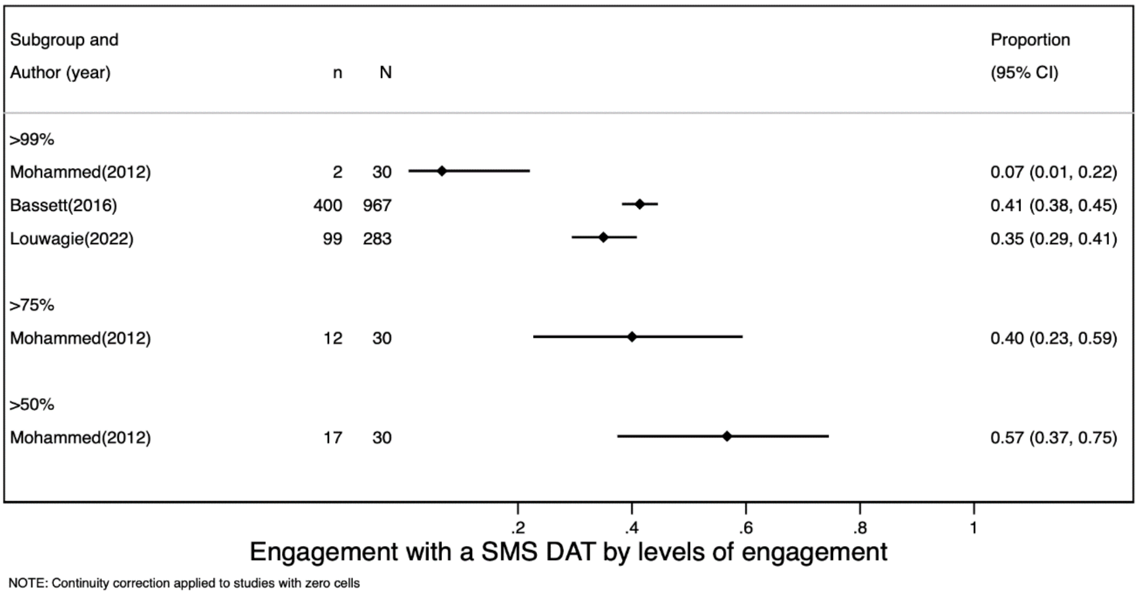

*Figure S6. Sustained engagement with a DAT by levels of engagement - Pillbox*

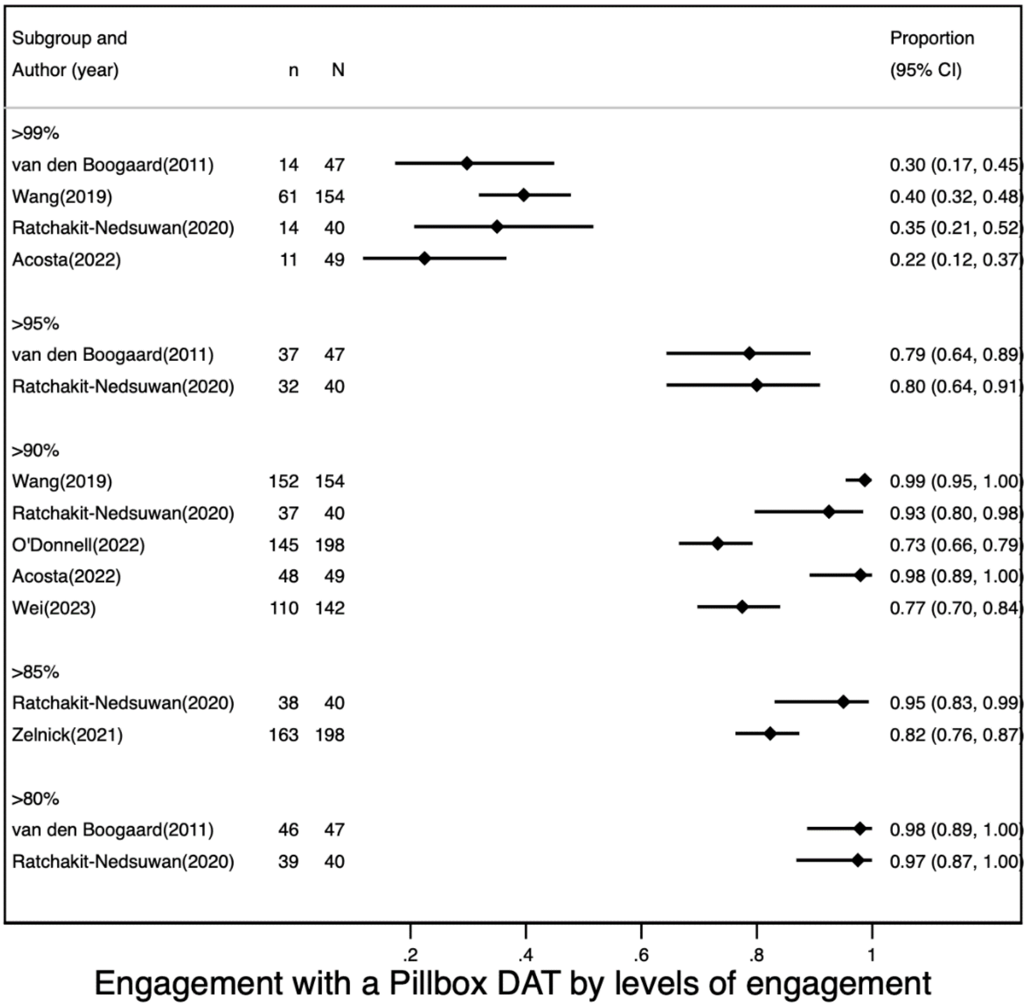

NOTE: Continuity correction applied to studies with zero cells

*Figure S7. Sustained engagement with a DAT by levels of engagement – 99DOTS*

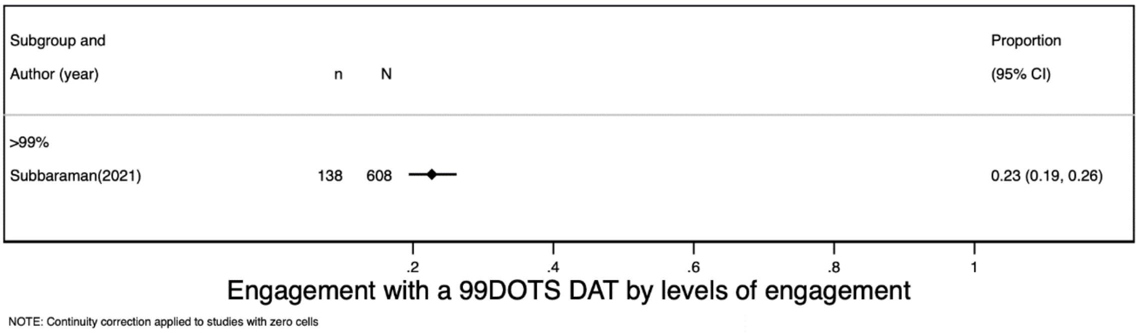

*Figure S8. Sustained engagement with a DAT by levels of engagement - VST*

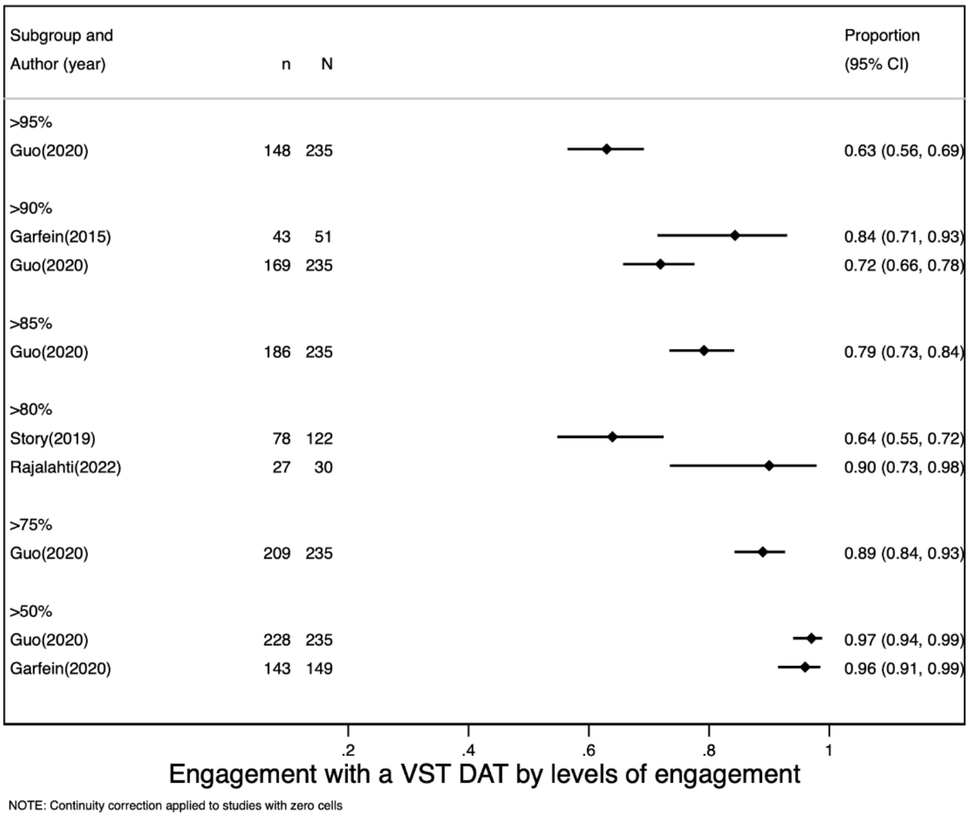

*Figure S9. Sustained engagement with a DAT by levels of engagement - Phone*

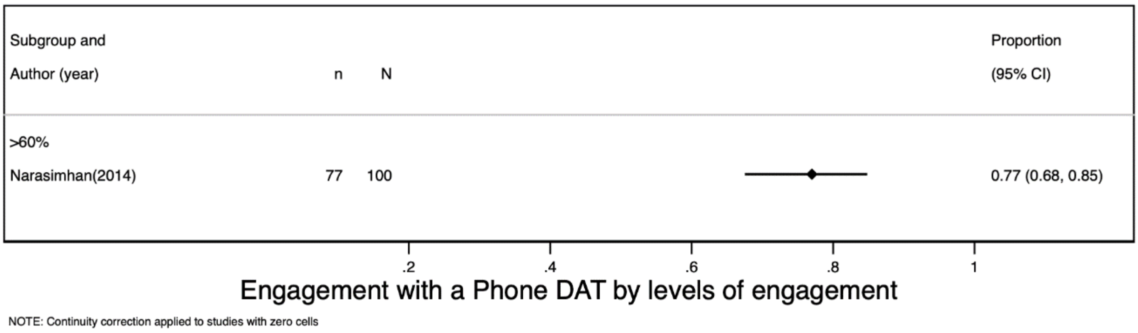

**Engagement by treatment phase**

Considering engagement by treatment phase, where measures of DAT engagement over time were presented, these are summarised in Table S7. We found very few studies that showed engagement over different treatment phases.

*Table S7. Varied measures of engagement with a DAT by treatment phase (Reach C)*

| Author | Year | Description of the measure | n | N | % |
|---|---|---|---|---|---|
| **Intensive phase** | | | | | |
| Moulding | 2002 | Proportion of individuals with >90% adherence in the first 11 weeks. | 74 | 106 | 70% |
| Moulding | 2002 | Proportion of individuals with >80% adherence in the first 11 weeks. | 92 | 106 | 87% |
| **Continuation phase** | | | | | |
| Moulding | 2002 | Proportion of individuals with >90% adherence in months 4-12. | 62 | 106 | 58% |
| Moulding | 2002 | Proportion of individuals with >80% adherence in months 4-12. | 88 | 106 | 83% |
| Menzies | 2005 | Proportion of patients who took >80% of doses within 20 weeks for 4RMP or 43 weeks for 9INH (latent tuberculosis tretament). | 86 | 104 | 83% |
| Louwagie | 2022 | Proportion of patients with optimal adherence (assessed using adherence index using recall) over the last 4 days at 3 months of treatment. | 319 | 348 | 92% |
| Louwagie | 2022 | Proportion of patients with optimal adherence (assessed using adherence index using recall) over the last 4 days at 6 months of treatment. | 120 | 348 | 34% |

*Table S8. Varied measures of sustained engagement (Reach C).*

| Author | Year | Description of numerator | Description of denominator | n | N | % |
|---|---|---|---|---|---|---|
| Acosta | 2022 | Number of doses that were after the scheduled hour. | total # doses (from 49 PWTB) | 586 | 2569 | 23% |

| | | | | | | |
|---|---|---|---|---|---|---|
| Bachina | 2022 | Number of doses observed by VST | total # of prescribed doses (from 49 PWTB) | 40 | 49 | 81% |
| Bachina | 2022 | PWTB who used VST post-COVID | total # PWTB post covid | 12 | 18 | 67% |
| Bachina | 2022 | PWTB who used VST pre-COVID | total # PWTB pre covid | 11 | 31 | 35% |
| Burhan | 2019 | Number of PWTB using the app at the end of the 3-month period. | # PWTB in the study | 10 | 45 | 22% |
| Casalme | 2022 | Number of PWTB who stayed on VST until end of treatment. | Number of PWTB in VST group | 67 | 110 | 61% |
| Chen | 2020 | Number of PWTB withdrawing from the programme. | # PWTB choosing VST | 4 | 80 | 5% |
| Cross | 2019 | Number of PWTB who made a call on day 167 or later. | Number of PWTB on treatment at day 167. | 13 | 20 | 65% |
| O'Donnell | 2022 | Number of PWTB with > 90% adherence. | Total number of PWTB. | 145 | 198 | 73% |
| Trajman | 2010 | Number of PWTB with complete and reliable pillbox records to analyze adherence. | Number of PWTB in the study without medication adverse effects who had their adherence analyzed. | 676 | 802 | 84% |
| Zelnick | 2021 | Number of PWTB with >85% adherence to Bedaquiline as measured by a DAT. | Number of PWTB enrolled. | 163 | 198 | 82% |
| Zhadnova | 2020 | Number of PWTB with sustained engagement with the DAT after 6 months. | | 15 | 44 | 34% |

PWTB person with TB ; VST video-supported therapy; DAT digital adherence technology

**Doi plot showing study asymmetry**

*Figure S11. Doi plot of asymmetry in effect estimates [restricted to threshold>=90%; 18 data points].*

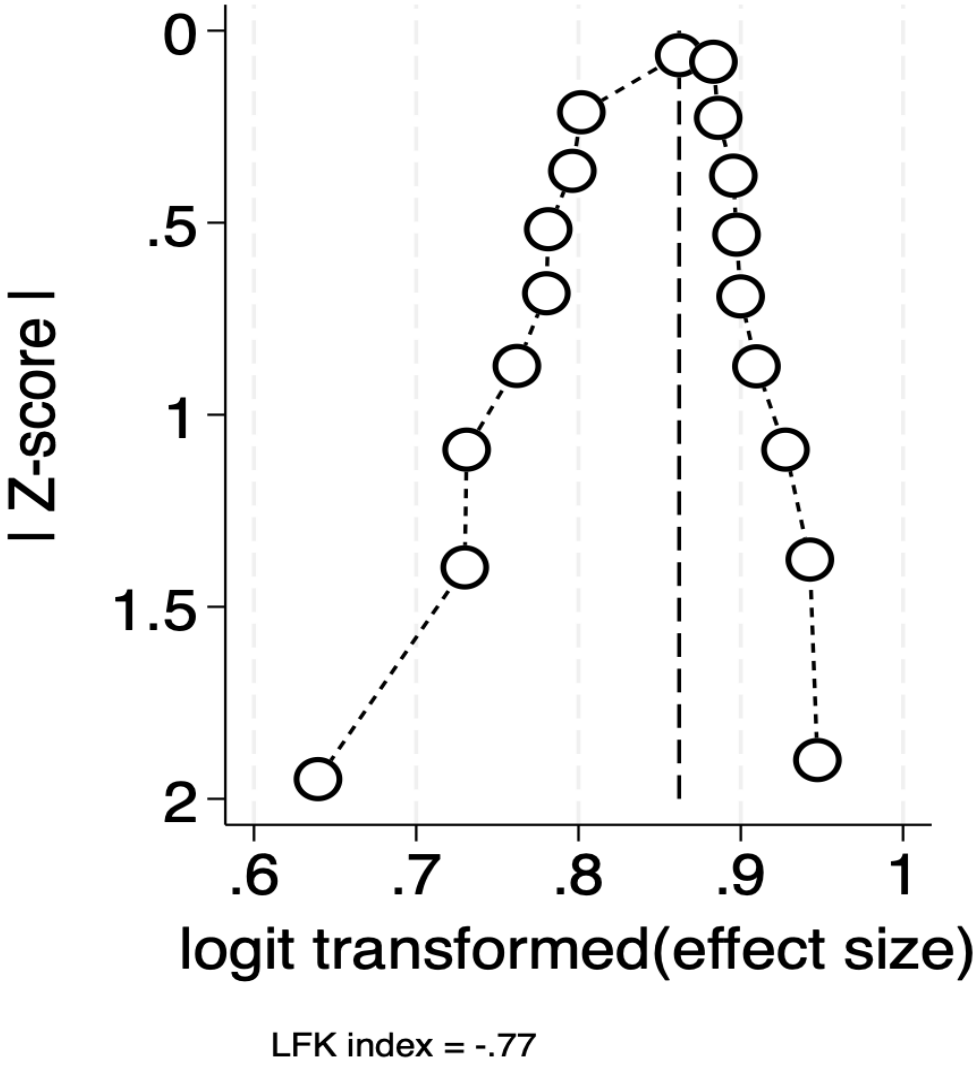

Doi plots and the Luis Furuya-Kanamori (LFK) index are alternatives to the funnel plot for assessing publication bias in a meta-analysis of proportions. Doi plots show normal quantile against effect size. When plotted, we can visually assess the symmetry (or assymmetry) in the effect estimates of studies, with each dot representing an individual study. Our a priori expectation is that studies showing higher levels of treatment adherence would be more likely to be published. Due to the high variability in measures included in this review, we assessed publication bias by evaluating similar non-engagement measures where the non-engagement threshold was >90% [18 data points]. When interpreting the LFK index, utilized to quantify asymmetry of small study effects from the Doi plot. With a LFK index less than +/-2 (-0.77) we do not find evidence of major asymmetry or publication bias [1,2].

*Table S9: Summary of technological challenges in DAT implementation*

| DAT type | Person/system | author (year) | Description | n/N | % | Additional notes |
|---|---|---|---|---|---|---|
| SMS | Person with TB | Hermans (2017) | % of persons with TB who had phone issues (stolen, not able to charge battery, damaged phone, no longer access to shared phone) | 45/171 | 26% | |
| | System | Gashu (2021) | % of persons with TB who received no messages | 4/152 | 3% | |
| | | Gashu (2021) | % SMS messages not delivered | | 15% | |
| | | Dessie Gashu (2020) | % of persons with TB who did not receive all reminders (over 2 months) | 3/23 | 13% | |
| | | Mohammed (2016) | % reminders not sent or received | 46276/220560 | 21% | |
| | | Hermans (2017) | % of persons with TB who did not receive all reminders | 16/171 | 9% | |
| | | Mohammed (2016) | of reminders not received, % due to system failures, GPRS (General Packet Radio Service) outage | | 67% | |
| Phone | Person with TB | Daftary (2017) | % of calls that failed due to PIN failure | 304/2867 | 11% | interactive voice response - messages sent, person with TB needed a PIN code to answer |
| | | Katende (2022) | % of voice messages not received by person with TB | 4/27 | 15% | Probably due to phone switched off |
| | System | Daftary (2017) | % of calls that failed due to network malfunction | 540/2867 | 19% | |
| | | Katende (2022) | % SMS not received by person with TB | 8/103 | 8% | (listed under phone as it was the main DAT component) |

| | | | | | | |
|---|---|---|---|---|---|---|
| 99DOTS | person with TB | Kiwanuka (2022) | % SMS received on handsets of people with TB, of all reminders sent | 58547/58831 | 99.50% | |
| | | Efo (2021) | % of persons with TB who could not call back at least once due to lack of phone credit | 58/200 | 29% | |
| | HCP | Kiwanuka (2022) | % SMS reminders sent among eligible days | 58831/65045 | 90.40% | |
| | | Guzman (2023) | % HCP who could not access adherence data due to no electricity, poor network, App not working (at any point) | 41/70 | 59% | |
| | | Guzman (2023) | % HCP who received DAT data daily | 56/61 | 92% | |
| | | Guzman (2023) | Of HCPs who cannot access adherence data, the % due to poor network connection | 34/41 | 83% | |
| | | Guzman (2023) | Of HCPs who cannot access adherence data, the % due to App not working | 33/41 | 80% | |
| | System | Kiwanuka (2022) | % days with SMS sending failure due to service outage | 29/357 | 8.12% | |
| Pillbox | Person with TB | Liu (2015) | % of all active pillbox problems due to incorrect pillbox use by person with TB | 429/1610 | 27% | |
| | | Liu (2015) | % of all active pillbox problems due to no power | 537/1610 | 33% | |
| | System | Liu (2015) | % of persons with TB who experienced any DAT problem (e.g., incorrect setting, lack of power, failure, other issues ) | 1003/2061 | 49% | reasons were multiple |
| | | Liu (2015) | % of all active pillbox problems for which the pillbox had to be replaced | 284/1610 | 18% | |
| | | Liu (2015) | % of all active pillbox problems due to pillbox failure | 440/1610 | 27% | |

| | | | | | |
|---|---|---|---|---|---|
| | Liu (2023) | % error days | 2812/193,200 | 1% | |
| | Trajman (2010) | % of persons with TB with incomplete pillbox data | 126/802 | 16% | reasons included loss/destruction of bottles; problems with the electronic device; repeat openings of box |
| | Wang (2020) | % doses with missing instances of pillbox data | 1526/6381 | 24% | Reasons for missing pillbox data include incorrect usage by HCP/person with TB; person with TB did not bring pillbox to clinic; person with TB did not attend visit |
| | Acosta (2022) | % of persons with TB who incorrectly did not receive SMS at least once | 9/49 | 18% | |
| | Acosta (2022) | % of persons with TB who incorrectly did receive SMS at least once | 37/49 | 76% | |
| | de Sumari-de Boer (2016) | of reminders sent, % that were incorrect (median across 5 persons with TB) | | 75% | |
| | de Sumari-de Boer (2016) | of reminder sent, % not received on phone | 46/500 | 9% | |
| HCP | Guzman (2023) | % HCP who could not access adherence data due to no electricity, poor network, App not working | 6/20 | 30% | |
| | Guzman (2023) | % HCP who received DAT data daily | 17/20 | 85% | |
| | Liu (2015) | % of all active pillbox problems due to doctor setting active pillbox incorrectly | 3/1610 | 0.2% | |
| | Liu (2015) | % of all active pillbox problems that were resolved | 1425/1610 | 89% | |
| | Guzman (2023) | Of those who cannot access adherence data, the % due to poor network connection | 4/6 | 67% | |

| | | | | | |
|---|---|---|---|---|---|
| | | Guzman (2023) | Of those who cannot access adherence data, the % due to App not working | 1/6 | 17% |
| VST | person with TB | Guo (2020) | % of persons with TB with problems uploading videos/app>10% of time | 26/235 | 11% |
| | | Chuck (2016) | % of persons with TB with technical problems using VST | 54/61 | 89% |
| | | Hoffman (2010) | % expected videos not received due to lost phone | | 10% |
| | | Nguyen (2017) | % of persons with TB with problems using VST platform | 16/40 | 40% |
| | | Garfein (2018) | % of persons with TB experiencing any problem with application | 132/214 | 62% |
| | | Garfein (2018) | % of persons with TB experiencing problem with application >half the time | 10/132 | 8% |
| | | Casalme (2022) | % persons with TB who stopped VST due to technology challenges | 7/22 | 32% |
| | | Chuck (2016) | of problems, % due to technical issues (overall) | 276/316 | 87% |
| | | Chuck (2016) | of problems, % due to person with TB-related smart-phone issues | 40/316 | 13% |
| | | Sekandi (2020) | of videos not transmitted, % due to phone issues | 616/919 | 67% |
| | | Sekandi (2020) | of videos not transmitted, % due to phone malfunction | 293/919 | 32% |
| | | Sekandi (2020) | of videos not transmitted, % due to phone battery issues | 223/919 | 24% |
| | | Sekandi (2020) | of videos not transmitted, % due to App issues | 97/919 | 11% |
| | | Sekandi (2020) | of videos not transmitted, % due to lack of internet | 48/919 | 5% |

| | | | | | | |
|---|---|---|---|---|---|---|
| | System | Garfein (2018) | % of persons with TB experiencing any cellphone reception issue | 147/212 | 69% | |
| | | Garfein (2018) | % of persons with TB experiencing cellphone reception issue> half the time | 20/147 | 14% | |
| | | Hoffman (2010) | % expected videos not received due to technical issues with transmission | | 25% | |
| | | Hoffman (2010) | % expected videos not received | | 50% | |
| | | Chuck (2016) | of all problems, % due to slow internet | 72/316 | 23% | |
| | | Sekandi (2020) | of videos not transmitted, % due to lack of internet | 48/919 | 5% | |
| | HCP | Guo (2020) | % HCP not satisfied with the quality of videos | 8/56 | 12% | |
| | | Guo (2020) | % HCP who experience problems with the app >10% of the time | 3/66 | 5% | |
| | | Bendiksen (2020) | % HCP who reported problems with video DAT | 5/17 | 29% | |
| | | Bendiksen (2020) | % days that video conferencing not undertaken due to technical issues | 268/3023 | 9% | unclear if from person with TB or HCP side or both |
| | | Krueger (2010) | % video calls where there were technical difficulties experienced | 73/4420 | 2% | unclear if from person with TB or HCW side or both |
| Ingestible sensor | | Belknap | % sensor-days where wearable component was not working | 34/1080 | 3.1% | |
| | | Belknap | % of sensors not identified correctly by the system (issue of wearable component/sensor functioning | 54/1080 | 5.0% | |

PWTB person with TB; HCP health care provider; SMS short message service; DAT digital adherence technology; PIN personal identification number; VST video-supported therapy

*Table S9: Summary of provider fidelity for DAT implementation*

| Author (year) | DAT (primary) | Description | n/N | % |
|---|---|---|---|---|
| Liu (2015) | SMS | % of participants who started intensive management, of those eligible | 40/265 | 15.1% |
| Liu (2015) | SMS | % of participants who started DOT management, of those eligible | 8/357 | 2.2% |
| Liu (2015) | pillbox | % of participants who started intensive management, of those eligible | 32/162 | 19.8% |
| Liu (2015) | pillbox | % of participants who started DOT management, of those eligible | 9/240 | 3.8% |
| Liu (2015) | SMS&pillbox | % of participants who started intensive management, of those eligible | 44/170 | 25.9% |
| Liu (2015) | SMS&pillbox | % of participants who started DOT management, of those eligible | 10/214 | 4.7% |
| Wang (2020) | pillbox | % of participants who started DOT management, of those eligible | 29/134 | 21.6% |
| Kiwanuka (2022) | 99DOTS | % of participants who reported their HCP contacted them, when eligible | 60/83 | 72.3% |
| Kiwanuka (2022) | 99DOTS | % of participants who received at least one type of support action among those eligible | 261/409 | 63.8% |
| Kumwichar (2022) | VST | % of dose-days when a reminder was sent, of eligible dose-days | 32/72 | 44.4% |
| Wei* (2023) | pillbox | % of participants who received VST, of those eligible - month 1 | 2/2 | 100.0% |
| Wei* (2023) | pillbox | % of participants who received VST, of those eligible - month 2 | 2/2 | 100.0% |
| Wei* (2023) | pillbox | % of participants who received VST, of those eligible - month 3 | 11/12 | 91.7% |
| Wei* (2023) | pillbox | % of participants who received VST, of those eligible - month 4 | 17/18 | 94.4% |
| Wei* (2023) | pillbox | % of participants who received VST, of those eligible - month 5 | 21/23 | 91.3% |
| Wei* (2023) | pillbox | % of participants who received VST, of those eligible - month 6 | 39/44 | 88.6% |
| Liu (2023) | pillbox | % of participants who were reported as receiving intensive management, of those eligible | 156/190 | 82.1% |
| Liu (2023) | pillbox | % of participants who were reported as receiving DOT management, of those eligible | 53/99 | 53.5% |
| Bassett (2016) | SMS | % of participants who received >=5 call attempts, as required by protocol | 694/967 | 71.8% |

SMS short message service; VST video-supported therapy; DOT directly observed therapy; HCP health care provider. "Participants" here reflects persons with TB

* Wei study was included as a preprint (2023), though these data were abstracted from the supplement of the subsequently published article (2024) [3,4].

*Table S10. Summary of included reports by DAT type, RE-AIM indicators, country income level, and calendar year by first author and publication year (n=105)*

| | HIC | | | | UMIC | | | | L/LMIC | | | |
|---|---|---|---|---|---|---|---|---|---|---|---|---|
| | **2000-2017** | | **2018-2023** | | **2000-2017** | | **2018-2023** | | **2000-2017** | | **2018-2023** | |
| | n | RE-AIM | n | RE-AIM | n | RE-AIM | n | RE-AIM | n | RE-AIM | n | RE-AIM |
| SMS | - | | 1 | Johnston (2018) | 3 | Bassett (2016); Lei (2013); Iribarren (2013) | 2 | Louwagie (2022); Khachadourian (2020) | 3 | Mohammed (2016); Mohammed (2012); Hermans (2017) | 3 | Cox (2018); Musiimenta (2020); Gashu (2021) |
| phone | 1 | Person (2011) | - | | - | | 5 | Iribarren (2022); Zhadnova (2020); I (2019); Li (2022); Saunders (2018) | 3 | Daftary (2017); Narasimhan (2014); Kittusami (2014) | 11 | Kalita (2021); Gashu (2021); Katende (2022); Santra (2021); Rao (2022); Ali (2019); Jose (2022); Mathur (2019); Kumar (2019); Ggita (2018); Gashu (2020) |
| 99DOTS | - | | - | | - | | - | | - | | 7 | Thakkar (2019); Cross (2019); Kiwanaku (2022); Cattamanchi (2021); Efo (2021); Subbaraman (2021); Thomas (2020) |
| Pillbox | 4 | Belknap (2017); Trajman (2010); Menzies (2005); Ailinger (2008) | 3 | Wei (2023); O'Donnell (2022); Scott (2023) | - | | 9 | Zelnick (2021); Ratchakit-Nedsuwan (2020); Wang (2019); Wang (2020); Guo (2021); Wei (2023); Liu (2023); Acosta (2022); Wang (2021) | 3 | de Sumari-de Boer (2016); Moulding (2002); van den Boogaard (2011) | 4 | Park (2019); Velen (2023); Muyindike (2022); Manyazewal (2022) |
| VST | 9 | Demaio (2001); Garfein (2015); Olano-Soler (2017); Krueger (2010); Chuck (2016); Mirsaedi (2015); Wade (2009); | 15 | Holzman (2018); Zhao (2022); Bachina (2022); Garfein (2020); Rajalahti (2022); Lam (2018); Lippincott (2022); Macaraig (2018); Story (2019); Bendiksen (2020); | 1 | Sinkou (2017) | 3 | Kumwichar (2022); Guo (2020); Peinado (2022) | 2 | Nguyen (2017); Hoffman (2010) | 4 | Casalme (2022); Holzman (2019); Sekandi (2021); Sekandi (2020) |

| | | | | | | | | | | | | |
|---|---|---|---|---|---|---|---|---|---|---|---|---|
| | | Wade (2012); Buchman (2017) | | Bommakanto (2020); Do (2019); Garfein (2018); Donahue (2021); Perry (2021) | | | | | | | | |
| Ingestion sensor | 1 | Belknap (2013) | 1 | Browne (2019) | - | | - | | - | | - | |
| Web-based Apps | - | | 1 | Kim (2021) | - | | 1 | Cocozza (2020) | 1 | Navin (2017) | 2 | Chen (2022); Adong (2022) |
| >1 DAT intervention | - | | - | | 1 | Liu (2015) | - | | - | | 2 | Guzman (2023); de Groot (2022) |

#### 2.2.2. References from which data were extracted for the scoping review

*The numbering does not relate to reference numbers in the main manuscript. 106 articles listed – 105 included in the review plus Wei 2024 for which the data from the supplement was used in the review.*

1) Cross A. 99DOTS: A low-cost approach to monitoring and improving medication adherence. ACM International Conference Proceeding Series. 2019.

2) Kim AJ, Yang J, Jang Y, Baek JS. Acceptance of an informational antituberculosis chatbot among Korean adults: mixed methods research. JMIR mHealth and uHealth. 2021;9: e26424.

3) Holzman SB, Zenilman A, Shah M. Advancing patient-centred care in tuberculosis management: a mixed-methods appraisal of video directly observed therapy. Open Forum Infect Dis. 2018;5.

4) DeMaio J, Schwartz L, Cooley P, Tice A. The application of telemedicine technology to a directly observed therapy program for tuberculosis: a pilot project. Clin Infect Dis. 2001;33: 2082–2084.

5) Cocozza AM, Linh NN, Nathavitharana RR, Ahmad U, Jaramillo E, Gargioni GEM, et al. An assessment of current tuberculosis patient care and support policies in high-burden countries. International Journal of Tuberculosis and Lung Disease. 2020;24: 36–42.

6) O'Donnell MR, Padayatchi N, Wolf A, Zelnick J, Daftary A, Orrell C, et al. Bedaquiline adherence measured by electronic dose monitoring predicts clinical outcomes in the treatment of patients with multidrug-resistant tuberculosis and HIV/AIDS. Journal of acquired immune deficiency syndromes (1999). 2022.

7) Do D, Garfein RS, Cuevas-Mota J, Collins K, Liu L. Change in Patient Comfort Using Mobile Phones Following the Use of an App to Monitor Tuberculosis Treatment Adherence: Longitudinal Study. JMIR MHealth UHealth. 2019;7: 11638.

8) Guo X. A comprehensive app that improves tuberculosis treatment management through video-observed therapy: usability study. JMIR Mhealth Uhealth. 2020;8.

15) Gashu KD, Gelaye KA, Lester R, Tilahun B. Effect of a phone reminder system on patient-centered tuberculosis treatment adherence among adults in Northwest Ethiopia: a randomised controlled trial. BMJ health & care informatics. 2021;28.

16) Johnston JC. The effect of text messaging on latent tuberculosis treatment adherence: a randomised controlled trial. Eur Respir J. 2018;51.

17) Wang XL, Lei J, Wang XW, Liu T, Lu JR, Tian XM. Construction and effect evaluation of tuberculosis information platform in Ningxia. Zhonghua yu fang yi xue za zhi [Chinese journal of preventive medicine]. 2021;55: 517–520.

18) Liu X. Effectiveness of electronic reminders to improve medication adherence in tuberculosis patients: a cluster randomised trial. PLoS Med. 2015;12.

32) Gashu KD, Gelaye KA, Tilahun B. Feasibility, acceptability and challenges of phone reminder system implementation for tuberculosis pill refilling and medication in Northwest Ethiopia. 2021.

33) Wade VA. Home videophones improve direct observation in tuberculosis treatment: a mixed methods evaluation. PLoS One. 2012;7.

34) Mohammed S, Glennerster R, Khan AJ. Impact of a daily SMS medication reminder system on tuberculosis treatment outcomes: a randomized controlled trial. Gao L, editor.

42) Kumar AA. Mobile health for tuberculosis management in South India: Is video-based directly observed treatment an acceptable alternative?'. JMIR mHealth and uHealth. 2019;7.

43) Navin K, Vadivu G, Maharaj A, Thomas T, Lavanya S, Engn, et al. A mobile health intervention to support TB eradication programme for adherence to treatment and a novel

QR code-based technique to monitor patient-DOTS provider interaction. 1st International Conference on Advanced Computational and Communication Paradigms (ICACCP). 2017;475: 41–54.

44) Musiimenta A, Tumuhimbise W, Atukunda EC, Mugaba AT, Muzoora C, Armstrong-Hough M, et al. Mobile health technologies may be acceptable tools for providing social support to tuberculosis patients in rural Uganda: A parallel mixed-method study. Tuberculosis Research & Treatment Print. 2020;2020: 7401045.

45) Ali AOA, Prins MH. Mobile health to improve adherence to tuberculosis treatment in Khartoum state, Sudan. J Public Health Africa. 2019;10.

46) Cox SN, Elf J, Lokhande R, Ogale YP, DiAndreth L, Dupuis E, et al. Mobile phone access and comfort: Implications for HIV and tuberculosis care in India and South Africa. Open Forum Infectious Diseases. 2018;5: S162.

47) Saunders MJ, Wingfield T, Tovar MA, Herlihy N, Rocha C, Zevallos K, et al. Mobile phone interventions for tuberculosis should ensure access to mobile phones to enhance equity - a prospective, observational cohort study in Peruvian shantytowns. Trop Med Int Health. 2018;23: 850–859.

48) Zhdanova SN, Ogarkov OB, Heysell SK. Mobile health intervention for outpatient treatment of Tuberculosis and HIV Infection. Acta biomedica scientifica. 2020;5: 46–53.

49) MaCaraig M, Lobato MN, McGinnis Pilote K, Wegener D. A national survey on the use of electronic directly observed therapy for treatment of tuberculosis. J Public Heal Manag Pract. 2018.

68) Iribarren S. TextTB: a mixed method pilot study evaluating acceptance, feasibility, and exploring initial efficacy of a text messaging intervention to support TB treatment adherence. Tuberc Res Treat. 2013;2013.

75) Lam CK, McGinnis Pilote K, Haque A, Burzynski J, Chuck C, Macaraig M. Using video technology to increase treatment completion for patients with latent tuberculosis infection on 3-month isoniazid and rifapentine: An implementation study. J Med Internet Res. 2018.

87) Rajalahti I, Kreivi HR, Ollgren J, Vasankari T. Asynchronous video supported treatment of tuberculosis is well adopted in a real-world setting - an observational study comparing two distinct applications. Infectious diseases (London, England). 2022; 04-Jan.

94) Chen A, Kumar R, Baria RK, Shridhar PK, Subbaraman R, Thies W. Impact of the 99DOTS digital adherence technology on tuberculosis treatment outcomes in North India: a pre-post study. 2022.
